# Associations between diet quality, epigenetic aging and epigenome: Findings from two population-based Studies

**DOI:** 10.1101/2025.05.17.25327830

**Authors:** Juliana F. Tavares, Dan Liu, Valentina Talevi, Fabian Eichelmann, Franziska Jannasch, Matthias B. Schulze, N. Ahmad Aziz, Ute Nöthlings, Monique M.B. Breteler

## Abstract

Several dietary patterns are suggested to benefit health, potentially through DNA methylation changes. However, to what extent adherence to so-called ‘healthy diets’ overlaps, whether these dietary patterns are equally beneficial, and whether they affect health outcomes through the same molecular mechanisms, remains unclear. Therefore, we investigated the overlap in adherence to ten diet quality scores, and examined the associations of these scores with both biological aging markers and DNA methylation profiles. We used data from the Rhineland Study, a large population-based cohort, and validated our findings using corresponding data from the independent EPIC-Potsdam cohort. Interestingly, we found minimal overlap of participants in the top 25% of adherence across different diet quality scores. Adherence to a healthy dietary pattern was associated with reduced epigenetic age acceleration regardless of the specific dietary pattern, except for the EAT-Lancet diet. Different dietary patterns were associated with distinct methylation profiles, which however largely converged onto the same biological pathways. Our research thus indicates that general adherence to a healthy dietary pattern promotes health through similar epigenetic mechanisms, despite variations in dietary composition.

## Introduction

With a staggering 11 million global deaths linked to unhealthy eating habits per year — outpacing the toll of smoking and other modifiable risk factors — improving diet quality stands as a cost-effective approach for promoting healthy aging and preventing chronic diseases.^1, 2^ However, much of the existing research in the field of nutrition tends to concentrate on individual foods and nutrients, lacking a comprehensive perspective on the overall health implications of dietary choices. A focus on dietary patterns, which encompass a wide range of nutrients, foods, and beverages, can offer a more cohesive perspective on how diet influences health outcomes.^3^ Commonly used methods to measure adherence to dietary patterns involve hypothesis-driven approaches, including diet quality scores, which measure to what extent an individual’s diet aligns with current nutritional knowledge or guidelines for disease prevention.^4, 5^ Several diet quality scores have been developed— for example, the Alternate Healthy Eating Index (AHEI),^6^ Mediterranean-style diet score (MDS),^7^ Dietary Approaches to Stop Hypertension (DASH),^8^ Mediterranean–DASH Intervention for Neurodegenerative Delay (MIND) diet,^9^ Nordic diet score^10^ and the Dietary Inflammatory Index (DII).^11^ These diet quality scores have been previously associated with a variety of chronic diseases and mortality.^12, 13, 14^ Additionally, plant-based diets as assessed by Plant-based Diet Indices (i.e. overall PDI, healthful PDI, and unhealthful PDI), and the EAT-Lancet diet, are receiving increasing attention because of their benefits to both human health and environmental sustainability.^15, 16, 17, 18, 19, 20^ Recent large-scale epidemiological studies investigating the association between eight dietary patterns and major chronic diseases as well as healthy aging parameters, found that adherence to a healthy diet was associated with a decreased risk of major chronic diseases and greater odds of healthy aging, although their relationship with specific health outcomes varied.^21, 22^ The extent to which adherence to various healthy dietary patterns overlaps, and whether they affect health outcomes through similar or distinct molecular mechanisms, has not been explicitly examined.

The molecular mechanisms through which diet quality affects various health outcomes remain largely unknown, yet DNA methylation changes have been suggested to be involved.^23, 24^ Epigenetic modifications to DNA represent a crucial molecular mechanism through which the environment including diet interacts with the genome.^25, 26^ Assessing the relationship between diet quality and the epigenome may thus reveal pathways underlying the biological mechanisms between diet quality and health outcomes. Indeed, recent work suggests that diet quality can significantly impact an individual’s epigenome,^27, 28^ which in part can be captured by epigenetic clocks that estimate biological age based on DNA methylation patterns.^29, 30, 31, 32, 33, 34^ Previous investigations have primarily linked individual food items and nutrients (i.e. red meat, fruits and vegetables, plasma carotenoids) with epigenetic clocks and the epigenome, yielding inconsistent results.^32, 33, 35, 36^ More recent studies have found that higher adherence to healthy dietary patterns correlated with slower biological aging.^37, 38^ In addition, a meta-analysis of epigenome-wide association studies (EWAS) of five US and European population-based cohorts using the Illumina 450K methylation array, which measures 450,000 CpG sites, showed that adherence to MDS and AHEI correlated with differential DNA methylation patterns across the epigenome,^27^ and that these diet-associated DNA methylation changes were associated with a lower risk of all-cause mortality.^27^ However, no studies have evaluated whether different dietary patterns (i.e. MDS, DASH, MIND, AHEI, Nordic, EAT-Lancet, PDI, hPDI, uPDI and DII) have distinct effects on biological aging and DNA methylation. Moreover, few EWAS of dietary patterns have been performed using the Human Infinium MethylationEPIC array that encompasses an unparalleled coverage of 850,000 CpGs sites across the genome. Importantly, whether diet-induced DNA methylation changes converge on distinct biological pathways that eventually contribute to various health outcomes remains unclear.

We investigated several commonly used, hypothesis-driven diet quality scores, with regard to their impact on the methylome and epigenetic markers of biological aging across the adult lifespan. We included both diet quality scores exclusively defined by their assumed beneficial effects on human health, and more recently proposed dietary patterns that aim to address human health as well as environmental sustainability (EAT-Lancet). Specifically, leveraging data from a large population-based cohort study in Germany, the Rhineland Study, we *1)* examined the relationship of ten hypothesis-driven diet quality scores with second-generation epigenetic indicators of biological age, *2)* performed EWAS studies on all these diet quality scores to identify differential DNA methylation profiles across the entire methylome, and *3)* conducted cross-omics functional analyses of diet-associated DNA methylation sites to reveal biological insights into the mechanisms underlying the effects of diet quality on health. Importantly, we replicated our analyses using corresponding data from the independent population-based EPIC-Potsdam cohort.

## Results

### Sample characteristics

The characteristics of the Rhineland Study sample are presented in **Table 1** and **Supplementary Table 1**. Of the 6,470 participants in our analytic sample, 3,665 (57%) were women. Participants’ mean (± standard deviation (SD)) age was 56·3 ± 13.6 years (age range: 30 – 95). In general, all diet quality scores were correlated with each other (**Figure 1a**): the uPDI and DII showed inverse correlations with all other indices but a positive association with each other (r = 0·61). Among all healthy dietary patterns, the Nordic diet and MDS scores had the strongest correlation (r = 0·58), while the Nordic diet and EAT-Lancet scores had the weakest correlation (r = 0·18). The epigenetic age acceleration estimates, GrimAA and PhenoAA, were positively, albeit weakly, correlated with each other (r = 0·22; **Suppl. Figure 2**). Only a small number of individuals consistently ranked within the top 25% for diet quality across all measured scores (n=18), or at least nine out of these scores (n=77; **Figure 1b**), with 1,212 participants in the top 25% adherence for at least one of the diet quality scores.

**Figure 1:**
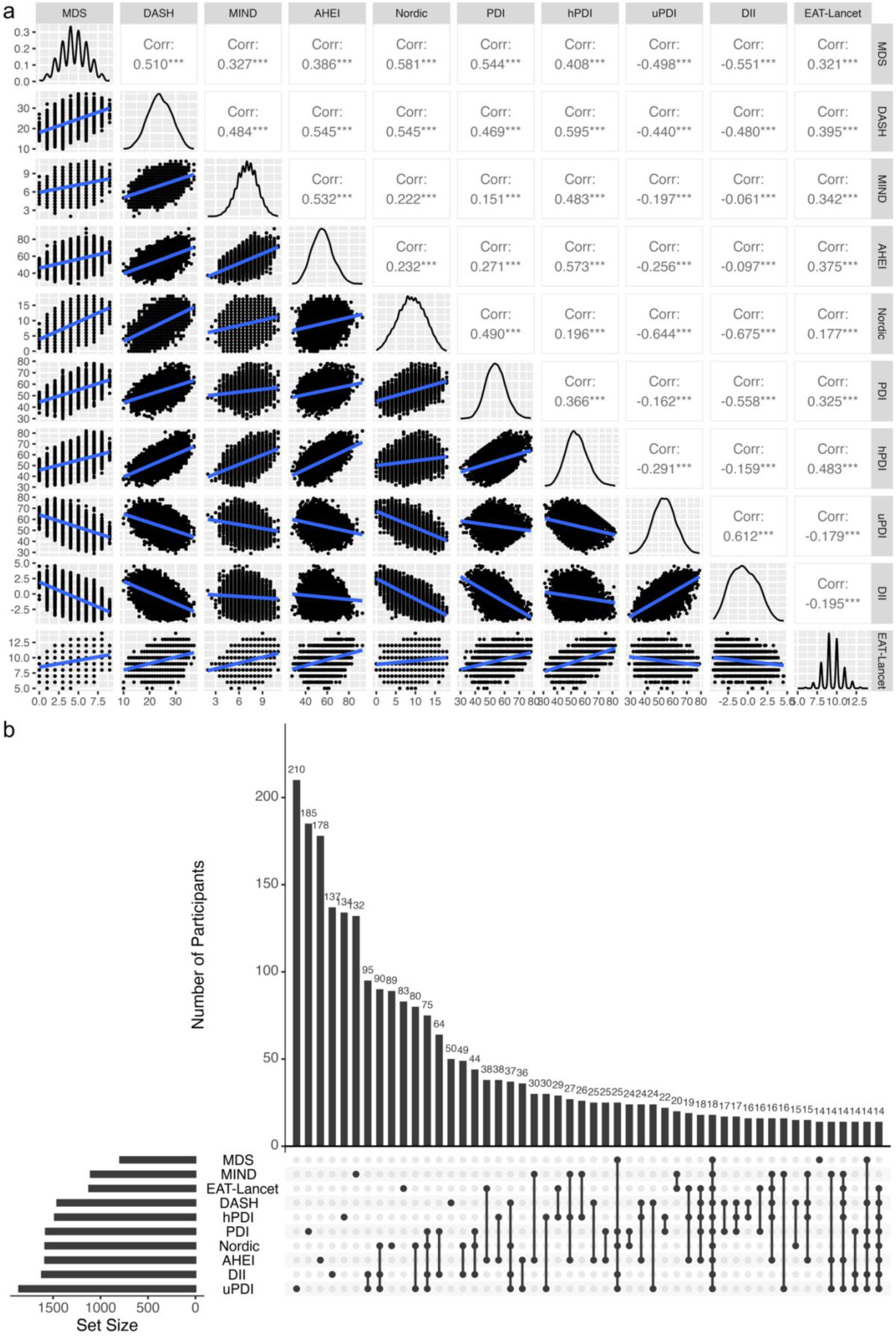
Correlation and overlap in high adherence (top 25%) across diet-quality scores in the Rhineland Study. A c*orrelation matrix with scatter plots, and density distribution of the diet-quality scores adherence* is depicted in Figure A. Correlation matrix plot of diet-quality scores is shown in the upper right triangle, with Pearson’s Correlation coefficient describing the relationship between each two scores. The significance code *** represent P<0.05. Scatter plots are shown in the lower left triangle and histograms of each diet-quality score are included in the diagonal. The UpSet plot (Figure B) summarize the overlap across the highest adherence (top 25%) across all the recommendation-based diet-quality scores. Bottom left horizontal bar graph represent the total number of participants within the highest quartile of adherence to healthy diets and lowest quartile of adherence to unhealthy diets across all ten recommendation-based diet quality score. The dots in each panel’s matrix represent unique and overlapping diet adherence. Connected dots designate a particular intersection between different diet-quality scores. The top bar graph in each panel recapitulates the number of participants for each unique recommendation-based diet-quality score or overlapping combination. Abbreviation: MDS, Mediterranean Diet Score; DASH, Dietary Approaches to Stop Hypertension; MIND, Mediterranean-DASH Intervention for Neurodegenerative Delay; AHEI, Alternate Healthy Eating Index score; PDI, Plant-based Diet Index; hPDI, Healthful Plant-based Diet Index; uPDI, Unhealthful Plant-based Diet Index; DII, Dietary Inflammatory Index

**Table 1:** Overview of the population characteristics ^1^.

| Characteristic |  | Rhineland Study<br>(n = 6470) |
| --- | --- | --- |
| Age, mean $y \pm SD$ | | 56.3 ± 13.6 |
| Women, n (%) |  | 3,665 (57) |
| Education ISCED11, n (%) | High | 3,400 (53) |
|  | Medium | 2,899 (45) |
|  | Low | 112 (2) |
| BMI, mean $\text{kg/m}^2 \pm SD$ | | 25.9 ± 4.5 |
| BMI, categories, n (%) | Normal weight | 2,964 (46) |
|  | Underweight | 73 (1) |
|  | Overweight | 3,050 (53) |
| Smoking status, n (%) | Never | 3,050 (47) |
|  | Former | 2,653 (41) |
|  | Current | 767 (12) |
| Total calories, mean $\text{kcal} \pm SD$ | | 2,489.2 ± 840.1 |
| Alcohol intake, mean $\text{g/day} \pm SD$ | | 19.5 ± 27.6 |
| MDS, mean score $\pm SD$ | | 4.5 ± 1.7 |
| DASH diet, mean score $\pm SD$ | | 24.0 ± 4.4 |
| MIND diet, mean score $\pm SD$ | | 7.0 ± 1.3 |
| AHEI-2010, mean score $\pm SD$ | | 55.5 ± 9.3 |
| Nordic diet, mean score $\pm SD$ | | 9.0 ± 3.4 |
| PDI, mean score $\pm SD$ | | 54.0 ± 6.7 |
| hPDI, mean score $\pm SD$ | | 54.0 ± 7.7 |
| uPDI, mean score $\pm SD$ | | 54.0 ± 7.8 |
| DII, mean score $\pm SD$ | | -0.5 ± 1.7 |
| EAT-Lancet, mean score $\pm SD$ | | 9.5 ± 1.2 |
| GrimAA, $y \pm SD$ | | -0.1 ± 7.3 |
| PhenoAA, $y \pm SD$ | | -0.1 ± 6.4 |

<sup>1</sup>Data were expressed as means ± SDs or numbers (percentages).
BMI categories: Underweight $<18.5 \text{ kg/m}^2$ ; Normal weight $18.5\text{-}24.9 \text{ kg/m}^2$ ; Overweight $\geq 25 \text{ kg/m}^2$ .
Recommendation based diet quality scores ranges: MDS 0-9; DASH 8-40; MIND 0-14; AHEI-2010 0-100; Nordic 0-18; PDI, hPDI and uPDI 18-90; EAT-Lancet 0-14.
Abbreviations: SD, standard deviation; MDS, Mediterranean Diet Score; DASH, Dietary Approaches to Stop Hypertension; MIND, Mediterranean-DASH Intervention for Neurodegenerative Delay; AHEI, Alternate Healthy Eating Index score; PDI, Plant-based Diet Index; hPDI, Healthful Plant-based Diet Index; uPDI, Unhealthful Plant-based Diet Index; DII, Dietary Inflammatory Index; BMI, body mass index; GrimAA, DNA methylation GrimAge Acceleration; PhenoAA, DNA methylation PhenoAge Acceleration.

### Association between diet quality and epigenetic age acceleration

In the Rhineland Study, higher adherence to the healthy dietary patterns including MDS, DASH, MIND, AHEI-2010, Nordic diet, PDI and hPDI tended to be associated with lower PhenoAA and GrimAA, while higher adherence to unhealthy dietary patterns (i.e. uPDI and DII) tended to be associated with higher PhenoAA and GrimAA (**Figure 2a**). Of note, the DASH, Nordic diet, and DII scores were consistently and significantly associated with both epigenetic aging metrics. Specifically, one SD increase in the DASH score (4.4 points) was associated with 0·23 years reduction in GrimAA (95% CI: −0·40 to −0·06) and 0·16 years in PhenoAA (95% CI: −0·29 to −0·03). Similarly, for the Nordic diet, each SD increase (3·4 points) was associated with 0·27 years decrease in GrimAA (95% CI: −0·46 to −0·08) and 0·20 years in PhenoAA (95% CI: −0·35 to −0·06). Conversely, one SD increase of DII (1·7 units) was associated with 0·16 years increase in GrimAA (95% CI: 0·02 to 0·31) and 0·15 years in PhenoAA (β 95% CI: 0·04 to 0·26). There was no association between EAT-Lancet diet score and epigenetic aging metrics (**Figure 2a)**. The analyses of the diet quality scores in quartiles of the distribution aligned with the results of the continuous analyses (**Suppl. Table 2**). Adjustment for body mass index (BMI) slightly attenuated the associations between the diet quality scores and epigenetic age acceleration metrics (**Suppl. Figure 3)**. Stratified analyses showed similar associations between diet quality and epigenetic aging metrics for both sexes (**Suppl. Figure 4**), and for people with and without obesity (**Suppl. Figure 5**). Interestingly, higher adherence to MDS, DASH, AHEI, Nordic, or PDI, and lower adherence to DII, was associated with lower GrimAA and PhenoAA among current smokers, but not in former or never smokers (**Figure 2b**).

**Figure 2:**
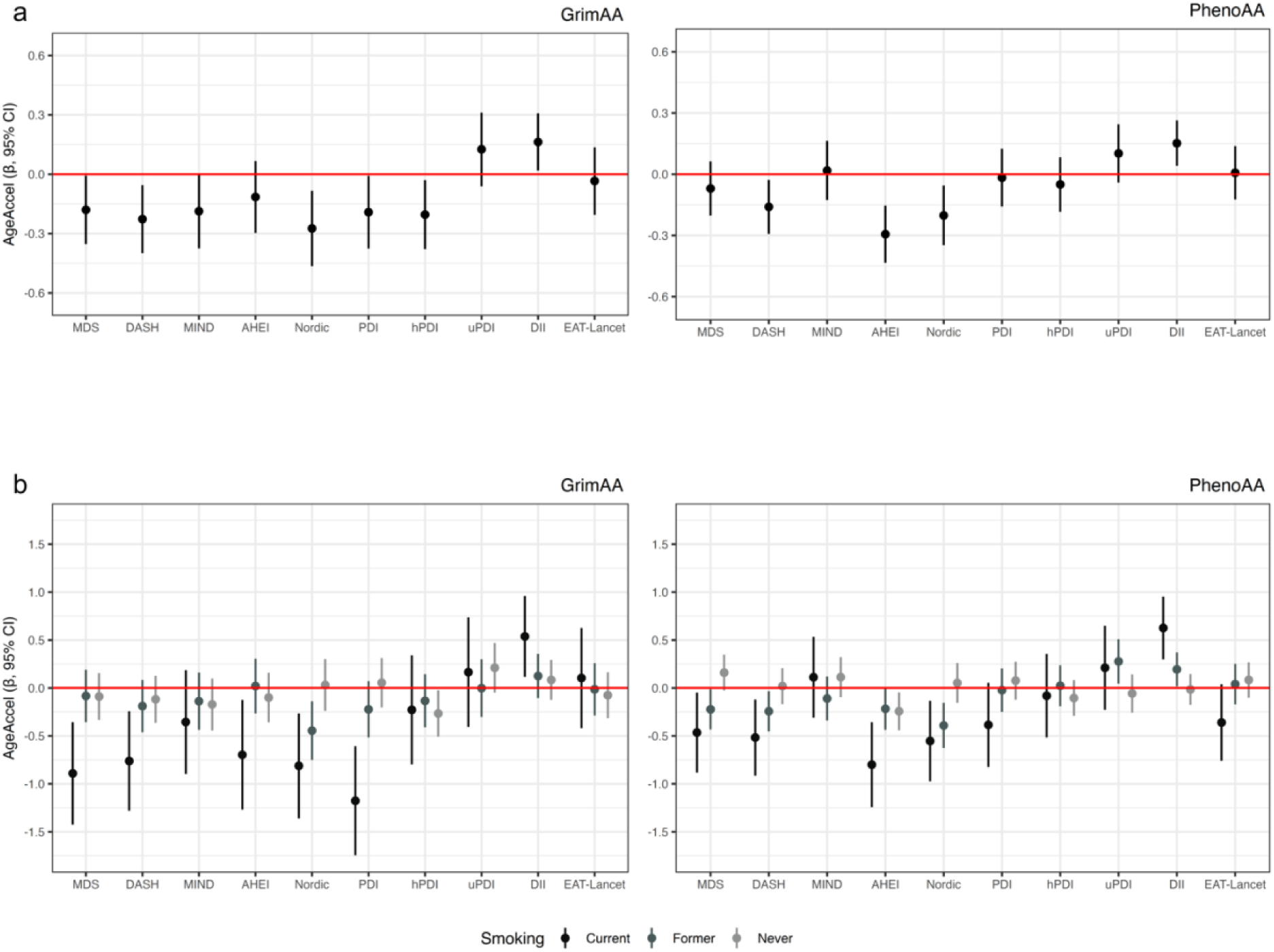
Associations between recommendation-based diet quality scores and two measures of epigenetic age acceleration in the Rhineland Study. The upper forest plots (Panel A) display the β-coefficients and 95% CIs from adjusted linear regression models, which represent the adjusted mean difference (in years) for the epigenetic acceleration metrics per 1-SD increase in diet quality (n = 6470). Models were adjusted for batch effect, blood cell proportion, sex, smoking status and total energy intake. Models testing associations with the DASH, Nordic, EAT-Lancet, PDI, hPDI and uPDI scores were additionally adjusted for alcohol intake (g/day). The lower forest plots (Panel B) display the β-coefficients and 95% CIs from adjusted linear regression models, which represent the adjusted mean difference (in years) for the epigenetic acceleration metrics per 1-SD increase in diet quality among never smokers (grey lines; n= 767), former smokers (dark grey lines; n= 2653), and never smokers (black lines; n= 3050). Models were adjusted for batch effect, blood cell proportion, sex and total energy intake. Models testing associations with the DASH, Nordic, EAT-Lancet, PDI, hPDI and uPDI scores were additionally adjusted for alcohol intake (g/day). Abbreviation: GrimAA, DNA methylation GrimAge Acceleration; PhenoAA, DNA methylation PhenoAge Acceleration; MDS, Mediterranean Diet Score; DASH, Dietary Approaches to Stop Hypertension; MIND, Mediterranean-DASH Intervention for Neurodegenerative Delay; AHEI, Alternate Healthy Eating Index score; PDI, Plant-based Diet Index; hPDI, Healthful Plant-based Diet Index; uPDI, Unhealthful Plant-based Diet Index; DII, Dietary Inflammatory Index

### Association between diet quality and epigenome

EWAS analyses revealed that except for EAT-Lancet all diet quality scores were associated with DNA methylation levels across the whole genome (**Figure 3**). The number of epigenome-wide significant CpG sites ranged from 54 for AHEI-2010 to 3 for uPDI (p < 5·9e-8) (**Suppl. Table 3**). Of note, half of the identified CpGs were only present on the methylationEPIC array, but not on the methylation450K array, and could thus not be identified in earlier studies. In general, healthy dietary patterns mostly exhibited negative β values. Consistently with this, for uPDI and DII we observed the opposite trend (**Suppl. Figure 6).**

**Figure 3:**
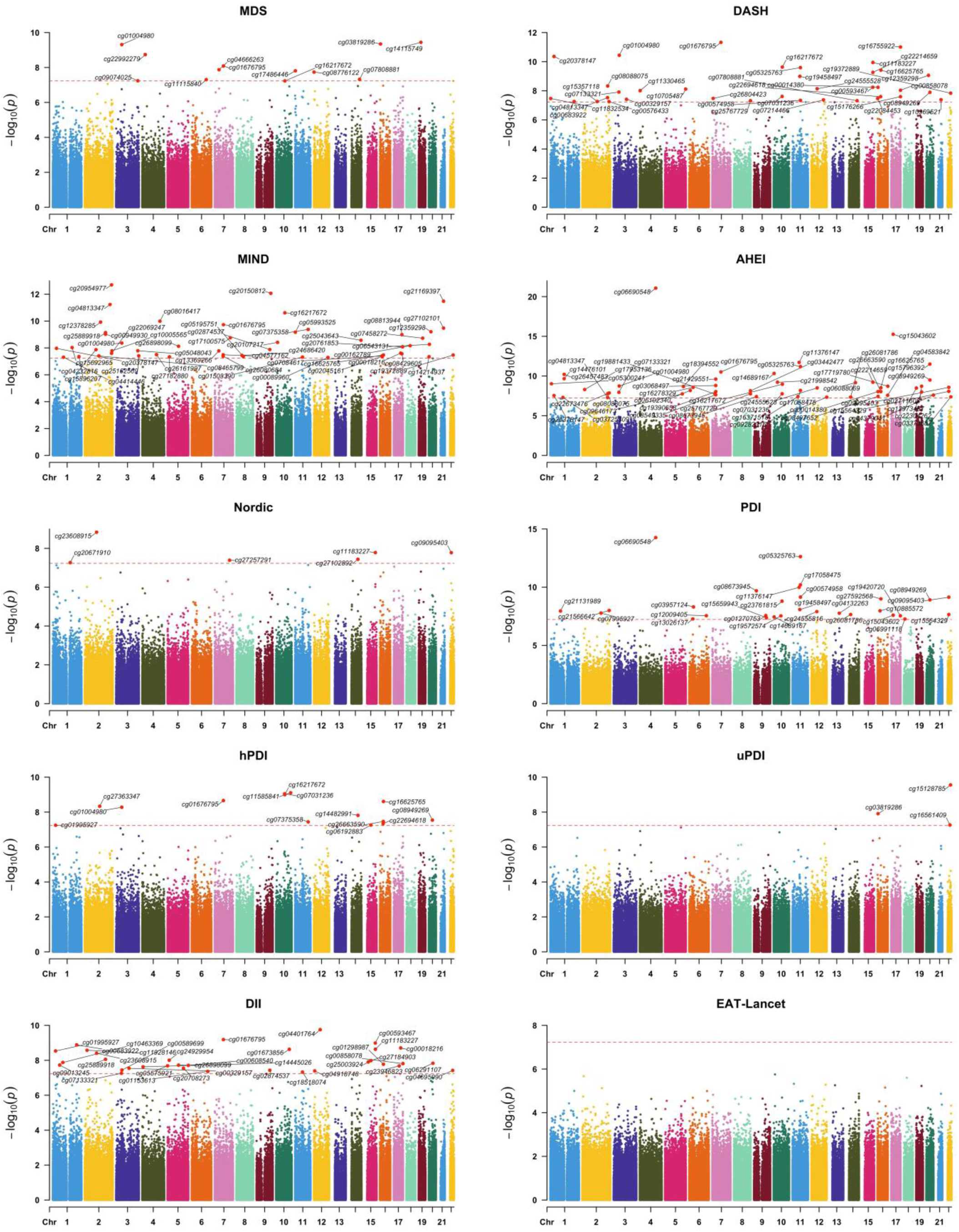
Manhattan plots of the epigenome-wide associations of diet quality scores in the Rhineland Study. Manhattan plots for each recommendation-based diet quality score. Results are plotted as negative log-transformed P values (y-axis) across the genome (x-axis). The red horizontal line represents the epigenome-wide significance based in Bonferroni threshold (5.95e-08, 0.05 divided by the number of CpGs). Linear models were adjusted for age, sex, batch effects, blood cell proportion, the first ten genetic principal components to account for population stratification, and smoking status. The top CpGs of each chromosome were annotated. Abbreviation: MDS, Mediterranean Diet Score; DASH, Dietary Approaches to Stop Hypertension; MIND, Mediterranean-DASH Intervention for Neurodegenerative Delay; AHEI, Alternate Healthy Eating Index score; PDI, Plant-based Diet Index; hPDI, Healthful Plant-based Diet Index; uPDI, Unhealthful Plant-based Diet Index; DII, Dietary Inflammatory Index.

### Validation of findings in an independent cohort

To validate our results, we conducted an independent replication analysis using data from 1,034 participants of the EPIC-Potsdam cohort. The mean age of the participants was 49.9 years (SD = 8.9), with 62% being women. Detailed sample characteristics are presented in **Supplementary Table 4**.

Overall, the associations between diet quality scores and both epigenetic age acceleration metrics in EPIC-Potsdam (**Suppl. Figure 7**) closely mirrored those observed in the Rhineland Study, both in direction and magnitude. Notably, the Nordic diet scores similarly exhibited significant negative associations with both PhenoAA and GrimAA. Specifically, one SD increase in the Nordic score was associated with 0·26 years reduction in GrimAA (95% CI: −0·42 to −0·09) and 0·30 years in PhenoAA (95% CI: −0·57 to −0·03). To further quantify the overall associations, we performed an inverse-variance weighted meta-analysis of results from the Rhineland Study and EPIC-Potsdam. The meta-analysis confirmed the patterns observed in each cohort independently, reinforcing the consistency of our findings. In particular, the pooled estimates for the DASH, Nordic, and DII scores remained statistically significant across both metrics of epigenetic age acceleration. The stratified analyses also mirrored the findings from the Rhineland Study, with no subgroup differences for BMI and sex (data not shown), but significant differences according to smoking status, where higher adherence to healthy dietary patterns and lower adherence to DII were especially associated with lower GrimAA and PhenoAA among current smokers (**Suppl. Figure 8**).

Of the 240 epigenome-wide significant CpGs identified in the Rhineland Study across nine dietary patterns, 39 were successfully replicated (p-value < 0.05 with consistent effect estimate directions). Among these, the uPDI (67%), AHEI (30%), and DASH (22%) dietary indices showed the highest replication rates (**Suppl. Figure 9**). Furthermore, even among CpGs that did not reach nominal significance (p > 0.05) in the replication cohort, more than 50% of the top CpGs retained consistent effect directions with the discovery cohort (**Suppl. Figure 9**&**10**). Beta-beta correlation analyses of the epigenome-wide significant CpGs (identified in the discovery cohort) further supported the consistency of the findings between the Rhineland Study and EPIC-Potsdam (r = 0.54, p < 0.001) (**Suppl. Figure 11**). Stratified analyses by diet quality score revealed varying degrees of correlation, with the highest reproducibility observed for CpGs associated with AHEI (r = 0.85), uPDI (r = 0.92), and DII (r = 0.67), while lower correlations were seen for CpGs linked to Nordic (r = 0.09) and MIND (r = 0.25) diets (**Suppl. Figure 12**).

No single CpG reached the epigenome-wide significance threshold (p < 5.95 × 10⁻⁸) for the EAT-Lancet diet score in either the discovery (Rhineland Study) or the replication (EPIC-Potsdam) cohort. Although we found CpGs associated with EAT-Lancet adherence in both cohorts at the suggestive significance level (p < 1 × 10⁻⁵), these completely differed between the two cohorts (**Suppl. Figure 13**), without a significant correlation between the beta values across CpGs (r = 0.007).

### Cross-omics functional analyses of diet-associated DNA methylation

In the Rhineland Study discovery cohort, we observed minimal overlap in the associated-CpGs (**Figure 4A**) and mapped genes (**Figure 4B**) across the 9 diet quality scores. In contrast, these distinct CpGs/mapped genes were involved in biological pathways that overlapped for more than 70% (**Figure 4C&D**, **Suppl. Figure 14**). These included the Kyoto Encyclopedia of Genes and Genomes (KEGG) pathways ‘adherens junction’, ‘axon guidance’, ‘calcium signaling pathway’, ‘endocytosis’, and the ‘MAPK signaling pathway’ (**Suppl. Table 9 & Suppl. Figure 15**), as well as the gene ontology (GO) pathways ‘actin binding’, ‘cell morphogenesis’, and ‘neurogenesis’ (**Suppl. Table 10 & Suppl. Figure 16**). Additionally, we identified diet quality scores-specific biological pathways that may underlie the distinct health effects of the corresponding diet. For instance, ‘aortic valve development and morphogenesis’, ‘heart valve development’ and ‘morphogenesis’ were uniquely associated with the DASH diet; whereas ‘cognition’, ‘learning or memory’ and ‘central nervous system neuron axonogenesis’ were uniquely associated with the MIND diet. Despite the minimal overlap in the diet-associated CpGs, the beta values of these CpGs were moderately correlated across diet scores (**Suppl. Figure 17**).

**Figure 4:**
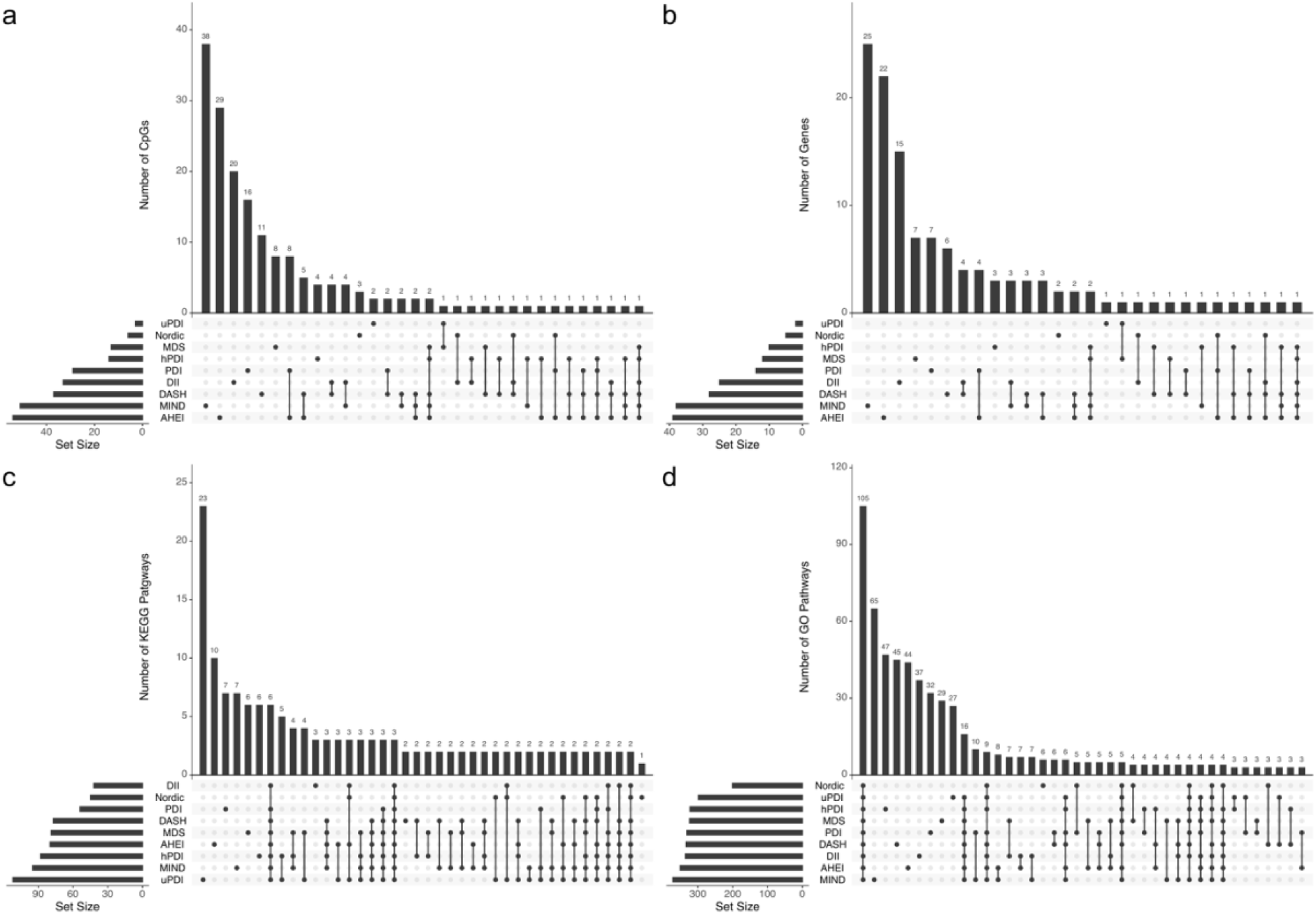
Overlap in diet-associated DNA methylation (epigenome-wide significantly associated CPGs), their mapped genes, and related KEGG and GO pathways, across different recommendation-based diet quality scores. The upper UpSet panels summarize the differentially methylated CpGs (panel A) and mapped genes (panel B) that overlap across the recommendation-based diet-quality scores. Bottom left horizontal bar graph represents the total number of CpGs or mapped genes for each recommendation-based diet quality score. The dots in each panel’s matrix represent unique and overlapping differentially methylated CpGs or its mapped genes. Connected dots designate a particular intersection between different groups of CpGs or its mapped genes. The top bar graph in each panel recapitulates the number of differentially methylated CpGs or mapped genes for each unique or overlapping combination. The lower UpSet panel summarize the number of KEGG (panel C) and GO (panel D) pathways that overlap across the recommendation-based diet-quality scores. Bottom left horizontal bar graph represents the total number of KEGG or GO pathways for each recommendation-based diet quality score. The dots in each panel’s matrix represent unique and overlapping KEGG or GO pathways. Connected dots designate a particular intersection between different groups of KEGG or GO pathways. The top bar graph in each panel recapitulates the number of KEGG or GO pathways for each unique recommendation-based diet-quality score or overlapping combinations. Abbreviation: MDS, Mediterranean Diet Score; DASH, Dietary Approaches to Stop Hypertension; MIND, Mediterranean-DASH Intervention for Neurodegenerative Delay; AHEI, Alternate Healthy Eating Index score; PDI, Plant-based Diet Index; hPDI, Healthful Plant-based Diet Index; uPDI, Unhealthful Plant-based Diet Index; DII, Dietary Inflammatory Index; KEGG, Kyoto Encyclopedia of Genes and Genomes; GO, Gene Ontology.

*In silico* analyses showed that several of the CpGs associated with MDS, DASH, MIND, AHEI, hPDI, and DII in our data, had been previously associated with AHEI in other cohorts (**Suppl. Table 5**). Moreover, these distinct dietary pattern-related CpGs have been previously linked to a wide range of diet-related traits (i.e., alcohol consumption, smoking), cardiometabolic traits (i.e., type II diabetes, BMI, waist circumference, CRP, insulin and glucose levels, blood pressure), lipid levels (i.e., HDL, LDL, triglycerides), protein levels as well as mortality (**Suppl. Table 5**). Interestingly, their mapped genes have been linked to several brain aging-related traits (including brain morphology, cortical surface area, subcortical volume and general cognitive ability (**Suppl. Table 5)**.

Diet-associated CpGs were associated with the expression levels of their mapped genes in both in *silico* analyses and our expression quantitative trait methylation (eQTM) analysis. For example, cg00574958 and cg17058475 (*CPT1A*), cg20954977 (*B3GNT7*), cg07458272 (*KIAA0355*), and cg02711608 (*SLC1A5*) were previously associated with the expression levels of the mapped genes in the BIOS database. Using individual-level gene expression data from the Rhineland Study, including expression data on 93 out of 126 mapped genes, we further extended these findings. We found that 33 diet-associated CpGs were associated with their mapped gene expression levels (**Figure 5**), including genes with important metabolic functions (i.e*., POR, SLC1A5, CPT1A*).^39^ Moreover, we observed that diet quality scores were associated with a small subset of these candidate genes, including *CPT1A, ABCA1*, *ATL3*, and *PFKFB2* (**Suppl. Figure 18**). Specifically, three distinct diet quality scores (i.e., DASH, AHEI and PDI) were related to *CPT1A* gene expression, thereby supporting the significance of this gene in metabolic pathways.

**Figure 5:**
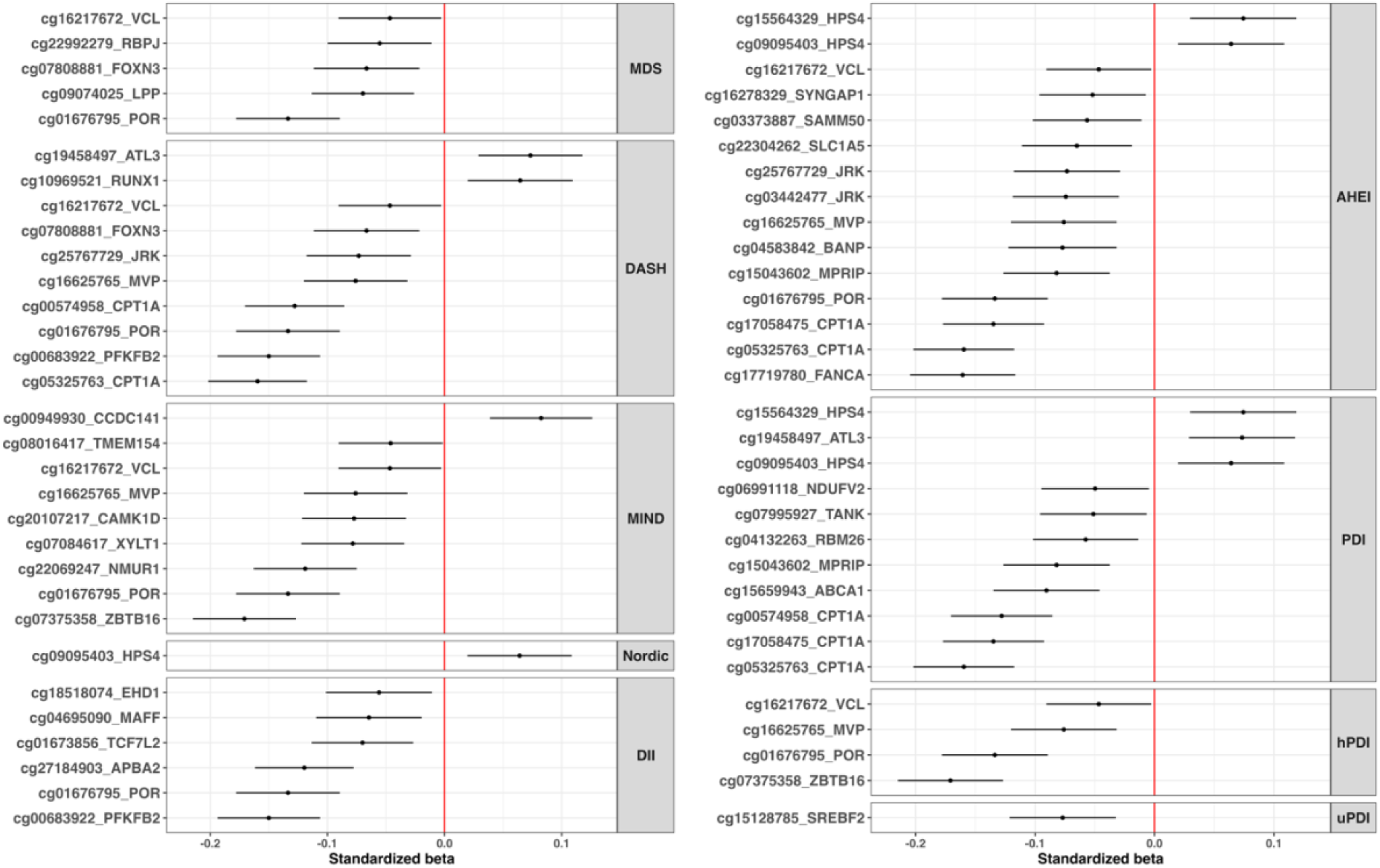
Diet-specific relations between associated CpGs and their mapped gene expression levels. Forest plots showing the effect estimate of CpGs on gene expression level, clustered by diet quality score. The standardized beta estimates are obtained through the following multivariable linear model: Gene_y_ ∼ intercept + CpG_x_ + age + sex + blood cell proportion + 10 PCs + smoking status + batch effects. Abbreviation: MDS, Mediterranean Diet Score; DASH, Dietary Approaches to Stop Hypertension; MIND, Mediterranean-DASH Intervention for Neurodegenerative Delay; AHEI, Alternate Healthy Eating Index score; PDI, Plant-based Diet Index; hPDI, Healthful Plant-based Diet Index; uPDI, Unhealthful Plant-based Diet Index; DII, Dietary Inflammatory Index.

Through the mQTLdb database, we uncovered several genetic variants (i.e., methylation quantitative trait loci (mQTLs)), which exhibited associations with the methylation levels of CpGs linked to the various dietary patterns. However, the majority of diet differential CpGs were not associated with any mQTLs. This is probably because the currently available mQTLs databases are all based on the 450K array, whereas half of the hits we identified are exclusive to the MethylationEPIC array.

## Discussion

Our results demonstrate that stronger adherence to healthier diets, as defined by multiple diet quality scores, barring the EAT-Lancet diet, was consistently associated with slower epigenetic aging in the general population. This trend was more prominent in current smokers. Intriguingly, different dietary patterns were associated with unique DNA methylation patterns, which however largely targeted similar biological pathways. These pathways predominantly involve processes related to cell structure, signaling, and metabolism, which are crucial in aging and chronic disease development. Additionally, we identified diet-specific pathways aligning with reported health claims for certain diets (e.g., DASH, MIND). Furthermore, these diet-associated CpGs have also been linked with aging-related traits, suggesting a functional connection between these epigenetic modifications and health outcomes.

Our observation that adherence to healthier diets was associated with slower biological aging aligns with findings from previous studies. Wang et al. found that adherence to a healthy diet, assessed through eight dietary patterns, is generally linked to a lower risk of major chronic diseases.^21^ More recently, Tessier et al. showed that higher adherence to healthy diets was associated with higher likelihood of healthy aging, as assessed according to measures of cognitive, physical and mental health, as well as living to 70 years of age free of chronic diseases.^22^ Kim et al. demonstrated a link between a higher DASH score and reduced epigenetic age acceleration.^37^ Similarly, Kresovich et al. reported an inverse relationship between four diet quality scores, including DASH, AHEI-2010 and a modified MDS, and epigenetic age acceleration in a cohort of non-Hispanic white females.^38^ Our study significantly extends those previous findings by not only reaffirming the connection between diet quality and markers of epigenetic aging acceleration, which focus on a limited number of CpG sites, but also by showing that the relationship between diet quality and the epigenome extends beyond these markers. Replication in the independent EPIC-Potsdam cohort further supported these findings by confirming a similar pattern of associations between diet quality scores and epigenetic aging indicators, along with partial replication at the epigenome-wide level. The stronger association between diet quality scores with epigenetic aging acceleration in current smokers, which was also replicated in the EPIC-Potsdam, indicates that better diet quality might mitigate the effects of other lifestyle factors associated with accelerated biological aging.

Our EWAS and cross-omics analyses in the discovery cohort illuminate the molecular mechanisms through which diet influences health outcomes. We found minimal overlap in the specific CpGs or genes associated with different diet quality scores, yet a substantial overlap in the associated biological pathways. Thus, while different healthy diets may influence distinct sets of epigenetic markers, they converge on similar biological processes. These shared pathways, including pathways related to cell signaling, metabolism, and neurogenesis, point to common molecular mechanisms underlying the link between nutrition and health. *In silico* analyses revealed that many diet-associated CpGs have been previously related to a wide range of health outcomes, supporting their relevance in mediating the health effects of diet. In addition, we identified diet-specific biological pathways that may underlie the specific health effects of the corresponding diet. For instance, the DASH diet has been associated with lower risk of CVDs, and exhibited methylation changes involved in heart function-related pathways. Similarly, MIND-related methylation changes were implicated in central nervous system axonogenesis and cognition-related pathways. From an applied perspective, the convergence of different dietary patterns on overlapping molecular pathways suggests that adopting a broadly healthy diet is likely beneficial at the population level. The fact that multiple, distinct dietary patterns share common epigenetic effects reinforces the public health message that promoting overall healthy eating patterns can have widespread health benefits. However, our finding that certain diet-specific pathways are also implicated—such as those related to cardiovascular health in the DASH diet or cognitive function in the MIND diet— highlights the potential for personalized dietary guidance. For individuals at higher risk of specific diseases, targeted dietary strategies could be developed based on the epigenetic markers and pathways most relevant to their risk profiles.

The variations in the strength of associations between different diet quality scores with epigenetic markers of biological aging and the methylome can be attributed to the structural differences in the conceptualization and calculation of these scores.^6–11,15,18^ While many of these scores harbor similarities, such as emphasizing the consumption of fruits, vegetables, and whole grains while limiting the intake of red and processed meats and sugars, they do diverge regarding specific food items or groups that are included or how they are accounted for (i.e., the same food item may be scored differently across different diets).^6, 7, 8, 9, 10, 11, 15, 18^ For instance, the MIND diet^9^ focuses on specific foods that contain nutrients that may support brain health—such as vitamin E, folate, n-3 fatty acids, carotenoids, and flavonoids— commonly found in berries, leafy greens, and moderate amounts of wine.

Similarly, the DASH diet also promotes plant-based foods but also prioritizes specific nutrient targets, such as sodium reduction, to support cardiovascular health.^8^ These differences may explain the limited overlap among participants with the highest adherence to various diet quality scores, and correspond to the distinct biological pathways associated with specific dietary patterns that may underlie possible diet-specific health effects, highlighting the potential for personalized dietary guidance. Nevertheless, we found comparable associations across different diet quality scores, and across the discovery and replication cohorts, supporting the generalizability of the beneficial effects of adherence to a general healthy diet.

Of note, we found little to no association of adherence to the EAT-Lancet diet with epigenetic aging acceleration or DNA methylation. We posit that this might stem from the EAT-Lancet diet’s recommended ranges, which emphasize environmental sustainability. Unlike the other diet quality scores, the EAT-Lancet sets upper intake limits, even for healthy food groups, and establishes a minimum intake of zero grams per day for several nutrient-dense food groups.^17, 18^ These intake ranges may not be sufficient to support adequate nutrient profiles to influence on epigenetic processes and pathways implicated in healthy aging, warranting further investigation in the context of aging research.

While our study provides substantial novel insights, it also has limitations. Our findings are based on a predominantly German population, and generalizability to persons with other ancestries or dietary habits remains to be investigated. Moreover, the FFQ-based dietary assessment is subject to measurement error and may not capture all dietary nuances. For example, they may not precisely capture specific or less frequently consumed foods. Despite these limitations, FFQs generally provide sufficient coverage of habitual dietary intake for assessing dietary patterns. Although multiple potential confounders were adjusted for in the present analysis, residual confounding could not be completely ruled out. Additionally, although a similar FFQ and DNA methylation assessment were employed between the discovery and replication cohorts, certain differences—particularly the fact that dietary data in EPIC-Potsdam were collected in the 1990s, possibly reflecting different eating patterns at the time, as well as potential regional variations in eating patterns despite both cohorts being from Germany—may account for some observed variance in effect estimates. Despite these nuances, the overall concordance in both direction and magnitude of associations supports their generalizability beyond the Rhineland Study population.

In conclusion, our results confirm the importance of promoting healthy dietary patterns as a crucial strategy for mitigating the aging process and its associated health risks. Additionally, our findings indicate that the choice of a specific healthy dietary pattern might not be as critical as the overarching adherence to a generally healthy diet. However, it is noteworthy that specific dietary patterns may offer additional benefits, potentially serving as tailored interventions for specific high-risk populations. Finally, the replication of our results in an independent cohort reinforces their generalizability, underscoring their potential impact on the development and implementation of public health programs aimed at promoting healthy aging.

## Methods

### Study Population

We used cross-sectional baseline data from the Rhineland Study, an ongoing population-based prospective cohort study. The study recruits individuals aged 30 years or older who live in either of two distinct municipal districts in Bonn, Germany, and who have a sufficient command of the German language to provide informed consent. A primary objective of the Rhineland Study is to identify determinants and markers of healthy ageing through a deep-phenotyping approach. At baseline, participants complete an 8-hour in-depth multi-domain phenotypic assessment, and various types of biomaterials (including blood, urine, stool, and hair) are collected. The ethics committee of the University of Bonn’s Medical Faculty approved the study, which is being conducted in accordance with the principles of the Declaration of Helsinki.

Between May 2016 and November 26, 2021, 8318 participants enrolled in the Rhineland Study. This study is based on the 6470 participants with complete information on diet, epigenetic data, and covariates of interest. Of these, 5768 participants with available genetic data were further selected for inclusion in our EWAS (**Supplementary** Figure 1).

### Dietary Intake and diet quality scores calculation

Participants’ dietary intake was assessed using a semi-quantitative Food Frequency Questionnaire (FFQ). The FFQ inquired about the consumption frequency of specified portion sizes of food and beverage items over the past year.^40^ For analysis inclusion, FFQs needed data on at least 80% of food items. Missing data were imputed with MissForest, as detailed elsewhere.^41^ Nutrient intake was estimated as the sum of nutrients content across all food items based on data from the German Food Code and Nutrient Database.^42^ The FFQ data served as the basis for estimating adherence to the following diet quality scores: MDS, DASH, MIND, AHEI-2010, Nordic, EAT-Lancet, PDI, hPDI, uPDI and DII.^6, 7, 8, 9, 10, 11, 15, 18^ For further details see **Supplementary Methods**.

### DNA methylation quantification

Genomic DNA was extracted from buffy coat fractions of anti-coagulated blood samples. DNA methylation levels were measured by Illumina iScan with Illumina’s Human MethylationEPIC BeadChip, which measures approximately 850,000 CpG sites across the genome. Detailed information is provided in the **Supplementary methods**.

### Estimation of epigenetic age acceleration

We calculated PhenoAge and GrimAge as epigenetic markers of biological age based on the algorithms developed by Levine et al. and Lu et al., using 513 and 1030 CpG sites, respectively.^32, 33^ The age acceleration estimators GrimAA and PhenoAA were defined as the residuals (in years) that result from regressing the PhenoAge and GrimAge age estimates on chronological age.

### Whole blood RNA isolation and gene expression profiling

PAXgene Blood RNA tubes (PreAnalytix/Qiagen) were used to collect fasting blood samples between 7:00 to 9:45 in the morning from an antecubital or dorsal hand vein. Total RNA was isolated according to manufacturer’s instructions using PAXgene Blood miRNA Kit and following the semi-automatic purification protocol (PreAnalytix/Qiagen). Gene expression sequencing was performed on the NovaSeq 6000 instrument (Illumina) platform. Detailed information on sequencing protocol and quantification of gene expression levels is provided in the **Supplementary methods**.

### Covariates

We used questionnaires and interviews to obtain information on participants’ characteristics including age, sex, history of medical conditions and medication, education and smoking status. Smoking status was defined as ‘current smoker’, ‘former smoker’ or ‘never’ based on self-report. Participants’ weight and height while in underwear by trained staff. Body mass index (BMI) was calculated as weight divided by height squared (kg/m^2^). Total daily energy intake (kcal/day) and alcohol consumption (grams/day) were derived from the FFQ.

### Statistical Analyses

The statistical analyses were conducted using R (version 4.2.1, The R Foundation). For sample demographics, we reported the mean and standard deviation (SD) for continuous variables, and the number and percentage for categorical variables. To compare the sample characteristics of the included and excluded participants, we performed binomial logistic regression adjusted for age and sex. The distributions of diet quality scores and age acceleration estimators were examined using histograms, and correlations among the diets were calculated using Pearson correlation coefficients. The overlap of participants in the top 25% for diet quality (highest quartile of MDS, DASH, MIND, AHEI-2010, Nordic Diet, EAT-Lancet, PDI and hPDI; and lowest quartile of uPDI and DII) across all diet quality scores, was illustrated using an UpSet plot.

### The association between dietary quality indices and epigenetic age acceleration

To investigate the relationship between dietary quality indices (independent variables) and two epigenetic age acceleration metrics (dependent variables: GrimAA, PhenoAA), we used multivariable linear regression models. Models were adjusted for sex, smoking status, total energy intake, batch effect and blood cell proportion estimated from the same DNA methylation samples.^43^ In the models examining associations with DASH, Nordic, PDI, hPDI, and uPDI scores, we additionally adjusted for alcohol intake (g/day). To evaluate the effect of BMI, we additionally adjusted for BMI in sensitivity analysis. In all analyses, the diet quality scores were transformed to have a mean of 0 and a standard deviation of 1. Therefore, all reported association estimates are based on a 1-standard deviation increase in diet quality. To determine whether the associations were primarily driven by individuals with the highest diet quality scores, we categorized the continuous diet quality scores in quartiles of their distributions. We then assessed linear trends by using the median value for each quartile category of the diet quality score as a continuous variable in our statistical model. We also evaluated effect modification of the diet quality scores by sex, smoking status, and BMI on epigenetic age acceleration with the addition of interaction terms in the models. In the interaction analysis, smoking status was considered as the 3 self-reported smoking categories (never, former, and current), and BMI was dichotomized into BMI <25 and ≥25 kg/m^2^.

Whenever an interaction term yielded a significant result (p ≤ 0.05), we performed a stratified analysis by the respective effect modifier. The effect estimates are presented with corresponding two-sided 95% confidence intervals. We considered p ≤ 0.05 as the threshold for statistical significance. To evaluate whether the model assumptions were met, we visually inspected the distribution of the residuals.

### Epigenome-wide association analysis (EWAS) of diet quality scores

We investigated the relationship between diet quality scores (independent variables) and DNA methylation level (dependent variable) across the genome using multiple linear regression, adjusting for age, sex, batch effects, blood cell proportion, the first ten genetic principal components to account for population stratification, smoking status and total energy intake (model 1). We further adjusted for BMI in model 2. We applied Bonferroni correction to account for multiple comparisons. An association was considered epigenome-wide significant if its p-value was smaller than the Bonferroni threshold (5.95e-08, 0.05 divided by the number of CpGs).

### Validation in an independent cohort

To externally validate our results, we conducted a replication analysis in an independent cohort, the EPIC-Potsdam study. This study employed similar methods for the measurement of dietary intake and DNA methylation levels as those used in the Rhineland Study. In EPIC-Potsdam, dietary intake was assessed using a semi-quantitative FFQ. The ten diet quality scores, ^6, 7, 8, 9, 10, 11, 15, 18^ MDS, DASH, MIND, AHEI-2010, Nordic, EAT-Lancet, PDI, hPDI, uPDI and DII, were calculated following the same methods as those in the Rhineland Study, as described in the Supplementary Methods. DNA methylation levels in participants were measured using Illumina’s Human MethylationEPIC BeadChip, while PhenoAge and GrimAge were calculated using the same established algorithms. The statistical analyses were applied identically to assess the associations between dietary quality indices and epigenetic age acceleration, as well as to perform an epigenome-wide association analysis (EWAS) of diet quality scores.

We conducted a meta-analysis using inverse-variance weighing to estimate the pooled association between dietary patterns and epigenetic age acceleration across cohorts. For each cohort, we obtained beta estimates and their corresponding 95% confidence intervals. The variance for each estimate was calculated as the square of the standard error. The pooled beta estimate was then computed as a weighted mean, where the weight for each cohort was the inverse of its variance. A 95% confidence interval for the pooled estimate was obtained using the standard error of the weighted mean. The meta-analyses was performed using the metafor package in R.

To confirm our EWAS findings, we examined whether the epigenome-wide significant CpGs identified in the discovery cohort (p-value < 5.95 × 10⁻⁸) were nominally significant (p-value < 0.05) and exhibited consistent effect estimate directions in the EPIC-Potsdam cohort. In addition, we evaluated the consistency of the associations by conducting a beta-beta correlation analysis between the beta estimates across all analyzed CpG sites in the two cohorts. Beta-beta plots and corresponding density plots were generated to visually inspect both the concordance of effect sizes and the overall distribution of beta estimates across cohorts. Furthermore, we specifically visualized the beta-beta correlations for the CpGs that reached epigenome-wide significance in the discovery cohort.

### Cross-omics functional analyses of diet-associated DNA methylation

In the discovery cohort, we explored the biological mechanisms underlying the effects of diet on the epigenome by examining diet-associated CpGs for enrichment in gene sets from the gene ontology database (GO) and KEGG pathway database using missMethyl R package. The analyses were performed for all diet-associated CpGs.

To explore whether the differential CpGs were associated with gene expression level, we explored expression quantitative trait methylation (eQTM) from the European blood-based BIOS QTL browser (https://molgenis26.gcc.rug.nl/downloads/biosqtlbrowser/) from BBMRI-NL. We further assessed whether the identified CpGs (independent variable) have an impact on their mapped gene expression levels in blood (dependent variable) using data from 1923 participants of the Rhineland Study for whom both methylation data and gene expression data were available. Linear regression models were adjusted for age, sex, batch effects, blood cell proportions, the first ten genetic PCs and smoking status. As a sensitivity analysis, we further assessed the associations between different diets and the genes mapped to diet-related CpGs. We used mQTLdb^44^ to examine whether there were previously identified genetic variants, the methylation quantitative trait loci (mQTLs), for the CpGs that were found to be associated with diet quality scores.

To identify whether the differential CpGs were associated with other traits, we looked up CpGs showing associations with the diet quality scores at epigenome-wide significance level using the EWAS Catalog (http://ewascatalog.org/). We also performed a look-up of known associations of the mapped gene for each CpG in previously published GWAS using the GWAS catalog (https://www.ebi.ac.uk/gwas).

## Data Sharing

The data used in this manuscript is not publicly available due to data protection regulations. Access to the Rhineland Study data can be provided to scientists in accordance with the study’s Data Use and Access Policy. Requests for additional information and/or access to the datasets can be sent to. Information on data access and contact details for the EPIC-Potsdam study can be obtained at https://www.dife.de/en/research/cooperations/epic-study/. All authors had full access to all the data in the study and take responsibility for the integrity of the data and the accuracy of the data analysis.

## Code availability

The code used to obtain results can be made available upon request.

## Acknowledgements

We would like to thank all participants of the Rhineland Study and the EPIC-Potsdam study, and the study personnel involved in data collection, processing, curation and management. The Rhineland Study is funded by the German Center for Neurodegenerative Diseases (DZNE). This work was supported through the Federal Ministry of Education and Research under the Diet-Body-Brain Competence Cluster in Nutrition Research (grant numbers 01EA1410C and 01EA1809C) and in the framework "PreBeDem - Mit Pravention und Behandlung gegen Demenz" (grant number 01KX2230), the Deutsche Forschungsgemeinschaft (DFG, German Research Foundation) under Germany’s Excellence Strategy (EXC 2151 – 390873048) and through SFB1454 – project number 432325352, and the Helmholtz Association under the 2023 and 2024 Innovation Pool. DL is partly supported by a grant from the Alzheimer’s Association (24AARFD-1192360). NAA is partly supported by a European Research Council Starting Grant (Number: 101041677). The work in EPIC-Potsdam was supported by a grant from the German Federal Ministry of Education and Research and the State of Brandenburg to the German Center for Diabetes Research (DZD; 82DZD00302 and 82DZD03D03).

## Contributions

DL, JFT and MMBB conceptualized and designed the study. VT, AA, and UN contributed to the study design and methodology. JFT, DL, FE, FJ and VT performed the statistical analysis and data visualization. All authors (JFT, DL, VT, AA, UN, FE, FJ, MBS, and MMBB) interpreted the results. JFT and DL prepared the initial draft of the manuscript. VT, AA and MMBB assisted in writing the manuscript. All authors critically reviewed and edited the manuscript. MBS (EPIC-Potsdam) and MMBB (Rhineland study) managed data curation and secured funding, and MMBB supervised the project.

## References

1. Astrup A, Dyerberg J, Selleck M, Stender S. Nutrition transition and its relationship to the development of obesity and related chronic diseases. Obes Rev 9 Suppl 1, 48–52 (2008).

2. Collaborators GBDD. Health effects of dietary risks in 195 countries, 1990-2017: a systematic analysis for the Global Burden of Disease Study 2017. Lancet 393, 1958–1972 (2019).

3. Hu FB. Dietary pattern analysis: a new direction in nutritional epidemiology. Curr Opin Lipidol 13, 3–9 (2002).

4. Ocke MC. Evaluation of methodologies for assessing the overall diet: dietary quality scores and dietary pattern analysis. Proc Nutr Soc 72, 191–199 (2013).

5. Zhao J, et al. A review of statistical methods for dietary pattern analysis. Nutr J 20, 37 (2021).

6. Chiuve SE, et al. Alternative dietary indices both strongly predict risk of chronic disease. J Nutr 142, 1009–1018 (2012).

7. Trichopoulou A, Costacou T, Bamia C, Trichopoulos D. Adherence to a Mediterranean diet and survival in a Greek population. N Engl J Med 348, 2599–2608 (2003).

8. Fung TT, Chiuve SE, McCullough ML, Rexrode KM, Logroscino G, Hu FB. Adherence to a DASH-style diet and risk of coronary heart disease and stroke in women. Arch Intern Med 168, 713–720 (2008).

9. Morris MC, et al. MIND diet slows cognitive decline with aging. Alzheimers Dement 11, 1015–1022 (2015).

10. Galbete C, et al. Nordic diet, Mediterranean diet, and the risk of chronic diseases: the EPIC-Potsdam study. BMC Med 16, 99 (2018).

11. Shivappa N, Steck SE, Hurley TG, Hussey JR, Hebert JR. Designing and developing a literature-derived, population-based dietary inflammatory index. Public Health Nutr 17, 1689–1696 (2014).

12. Reedy J, et al. Higher diet quality is associated with decreased risk of all-cause, cardiovascular disease, and cancer mortality among older adults. J Nutr 144, 881–889 (2014).

13. Harmon BE, et al. Associations of key diet-quality indexes with mortality in the Multiethnic Cohort: the Dietary Patterns Methods Project. Am J Clin Nutr 101, 587–597 (2015).

14. Sotos-Prieto M, et al. Association of Changes in Diet Quality with Total and Cause-Specific Mortality. N Engl J Med 377, 143–153 (2017).

15. Satija A, et al. Plant-Based Dietary Patterns and Incidence of Type 2 Diabetes in US Men and Women: Results from Three Prospective Cohort Studies. PLoS Med 13, e1002039 (2016).

16. Satija A, et al. Healthful and Unhealthful Plant-Based Diets and the Risk of Coronary Heart Disease in U.S. Adults. J Am Coll Cardiol 70, 411–422 (2017).

17. Willett W, et al. Food in the Anthropocene: the EAT-Lancet Commission on healthy diets from sustainable food systems. Lancet 393, 447–492 (2019).

18. Knuppel A, Papier K, Key TJ, Travis RC. EAT-Lancet score and major health outcomes: the EPIC-Oxford study. Lancet 394, 213–214 (2019).

19. Gibbs J, Cappuccio FP. Plant-Based Dietary Patterns for Human and Planetary Health. Nutrients 14, (2022).

20. Montejano Vallejo R, Schulz CA, van de Locht K, Oluwagbemigun K, Alexy U, Nothlings U. Associations of Adherence to a Dietary Index Based on the EAT-Lancet Reference Diet with Nutritional, Anthropometric, and Ecological Sustainability Parameters: Results from the German DONALD Cohort Study. J Nutr 152, 1763–1772 (2022).

21. Wang P, et al. Optimal dietary patterns for prevention of chronic disease. Nat Med 29, 719–728 (2023).

22. Tessier A-J, et al. Optimal dietary patterns for healthy aging. Nature Medicine, (2025).

23. Younesian S, Mohammadi MH, Younesian O, Momeny M, Ghaffari SH, Bashash D. DNA methylation in human diseases. Heliyon 10, e32366 (2024).

24. Zhang W, Song M, Qu J, Liu GH. Epigenetic Modifications in Cardiovascular Aging and Diseases. Circ Res 123, 773–786 (2018).

25. Bui H, et al. Association analysis between an epigenetic alcohol risk score and blood pressure. Clin Epigenetics 16, 149 (2024).

26. Fox FAU, Liu D, Breteler MMB, Aziz NA. Physical activity is associated with slower epigenetic ageing-Findings from the Rhineland study. Aging Cell 22, e13828 (2023).

27. Ma J, et al. Whole Blood DNA Methylation Signatures of Diet Are Associated With Cardiovascular Disease Risk Factors and All-Cause Mortality. Circ Genom Precis Med 13, e002766 (2020).

28. Do WL, et al. Epigenome-wide association study of diet quality in the Women’s Health Initiative and TwinsUK cohort. Int J Epidemiol 50, 675–684 (2021).

29. Bell CG, et al. DNA methylation aging clocks: challenges and recommendations. Genome Biol 20, 249 (2019).

30. Hannum G, et al. Genome-wide methylation profiles reveal quantitative views of human aging rates. Mol Cell 49, 359–367 (2013).

31. Horvath S. DNA methylation age of human tissues and cell types. Genome Biol 14, R115 (2013).

32. Levine ME, et al. An epigenetic biomarker of aging for lifespan and healthspan. Aging (Albany NY) 10, 573–591 (2018).

33. Lu AT, et al. DNA methylation GrimAge strongly predicts lifespan and healthspan. Aging (Albany NY) 11, 303–327 (2019).

34. Liu D, Aziz NA, Pehlivan G, Breteler MMB. Cardiovascular correlates of epigenetic aging across the adult lifespan: a population-based study. Geroscience 45, 1605–1618 (2023).

35. Quach A, et al. Epigenetic clock analysis of diet, exercise, education, and lifestyle factors. Aging (Albany NY) 9, 419–446 (2017).

36. Dugue PA, et al. Association of DNA Methylation-Based Biological Age With Health Risk Factors and Overall and Cause-Specific Mortality. Am J Epidemiol 187, 529–538 (2018).

37. Kim Y, et al. Higher diet quality relates to decelerated epigenetic aging. Am J Clin Nutr 115, 163–170 (2022).

38. Kresovich JK, Park YM, Keller JA, Sandler DP, Taylor JA. Healthy eating patterns and epigenetic measures of biological age. Am J Clin Nutr 115, 171–179 (2022).

39. Lai CQ, et al. Carbohydrate and fat intake associated with risk of metabolic diseases through epigenetics of CPT1A. Am J Clin Nutr 112, 1200–1211 (2020).

40. Nothlings U, Hoffmann K, Bergmann MM, Boeing H. Fitting portion sizes in a self-administered food frequency questionnaire. J Nutr 137, 2781–2786 (2007).

41. Richter R, Tavares JF, Miloschewski A, Breteler MMB, Mukherjee S. ImputeBench: Benchmarking Single Imputation Methods. medRxiv, 2025.2002.2002.25321536 (2025).

42. Dehne LI, Klemm C, Henseler G, Hermann-Kunz E. The German Food Code and Nutrient Data Base (BLS II.2). Eur J Epidemiol 15, 355–359 (1999).

43. Houseman EA, et al. DNA methylation arrays as surrogate measures of cell mixture distribution. BMC Bioinformatics 13, 86 (2012).

44. Min JL, et al. Genomic and phenotypic insights from an atlas of genetic effects on DNA methylation. Nat Genet 53, 1311–1321 (2021).

