## Supplementary Figures and Methods for "Associations between diet quality, epigenetic aging and epigenome: Findings from two population-based Studies"

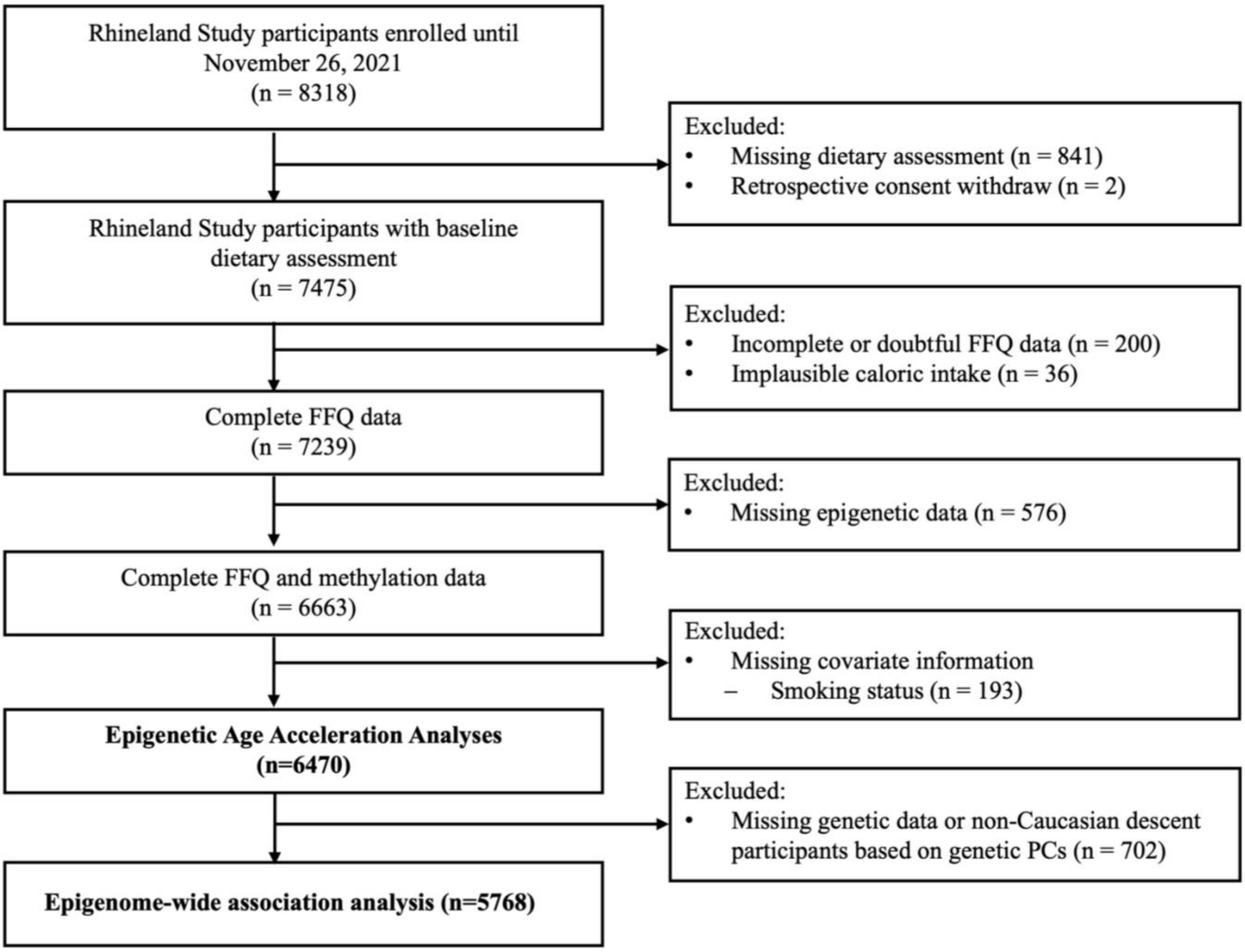

**Supplementary** ***Figure 1: Flow chart of participant selection overview.***

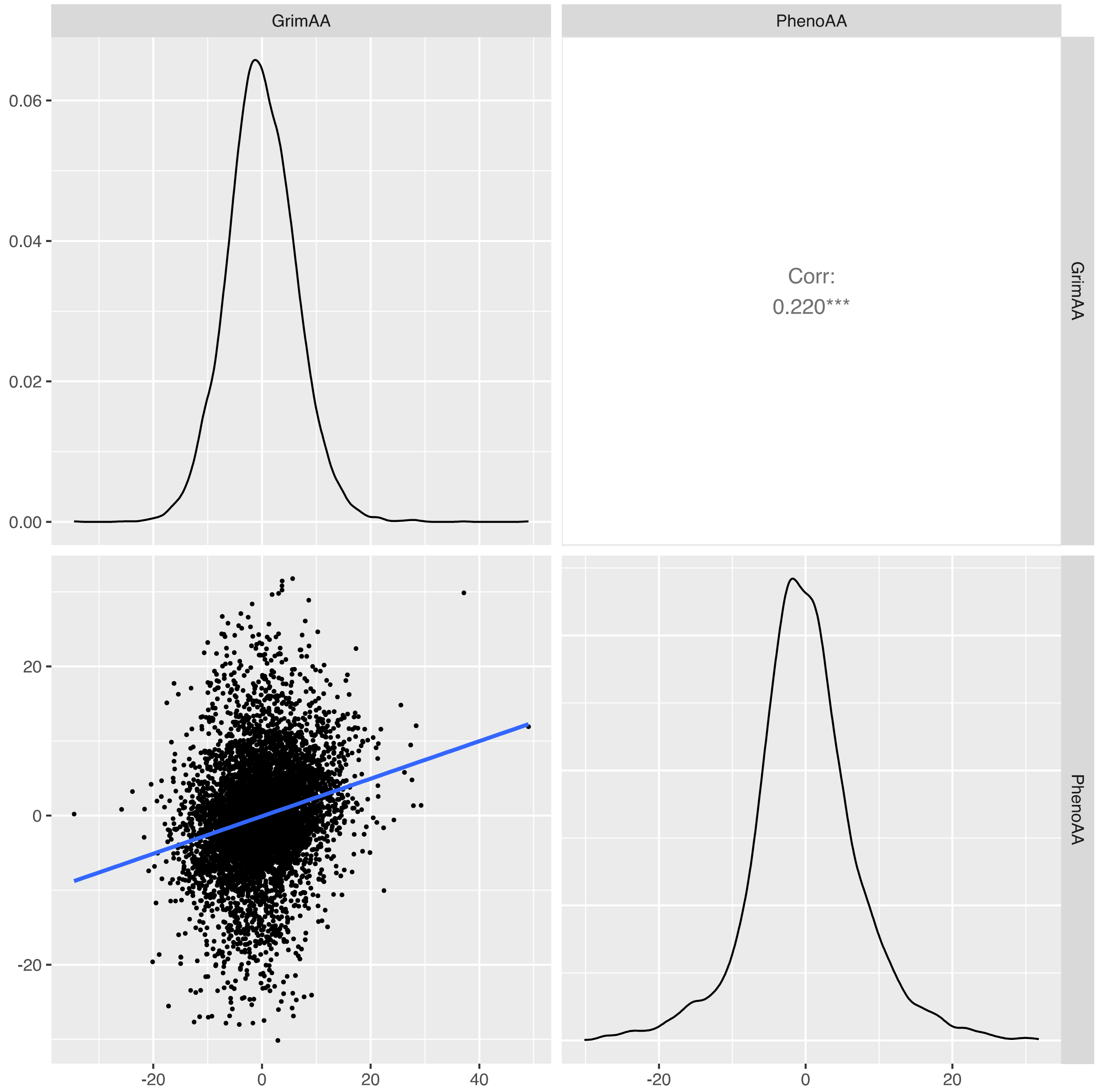

**Supplementary Figure 2.** ***Correlation matrix with scatter plot, and density distribution of the epigenetic age acceleration estimates.***

Correlation estimate of epigenetic age acceleration estimates is shown in the upper right figure, with Pearson's Correlation coefficient describing the relationship between the two estimates. The significance code *** represent P<0.05. The scatter plot is shown in the lower left figure and histograms of each epigenetic age acceleration estimate are included in the diagonal.

Abbreviation: GrimAA, DNA methylation GrimAge Acceleration; PhenoAA, DNA methylation PhenoAge Acceleration

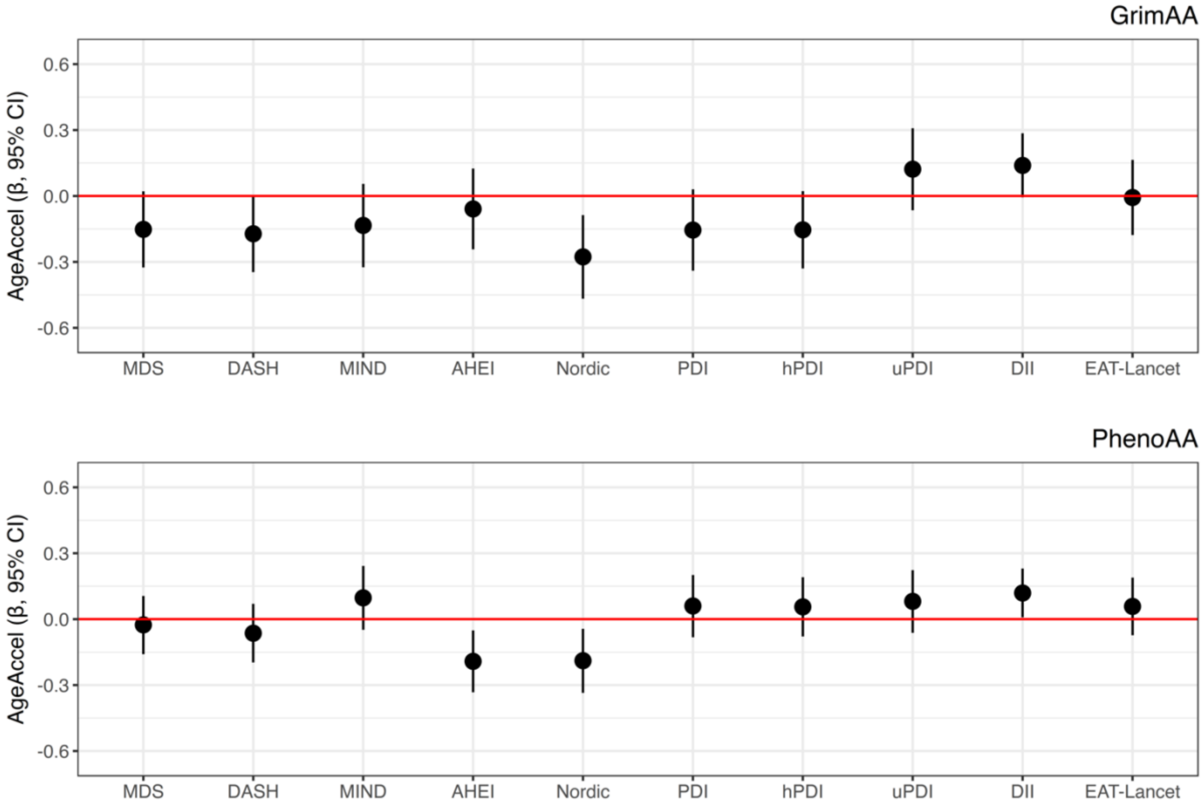

***Supplementary Figure 3: Associations between recommendation-based diet quality scores and the two measures of epigenetic age acceleration with additional model adjustment for body mass index.***

Plots display the β-coefficients and 95% CIs from adjusted linear regression models, which represent the adjusted mean difference (in years) for the epigenetic acceleration metrics per 1-SD increase in diet quality (n = 6435). Thirty-five participants were missing information on body mass index. Models were adjusted for batch effect, blood cell proportion, sex, smoking status, total energy intake and body mass index. Models testing associations with the DASH, Nordic, EAT-Lancet, PDI, hPDI and uPDI scores were additionally adjusted for alcohol intake (g/day).

Abbreviation: GrimAA, DNA methylation GrimAge Acceleration; PhenoAA, DNA methylation PhenoAge Acceleration; MDS, Mediterranean Diet Score; DASH, Dietary Approaches to Stop Hypertension; MIND, Mediterranean-DASH Intervention for Neurodegenerative Delay; AHEI, Alternate Healthy Eating Index score; PDI, Plant-based Diet Index; hPDI, Healthful Plant-based Diet Index; uPDI, Unhealthful Plant-based Diet Index; DII, Dietary Inflammatory Index.

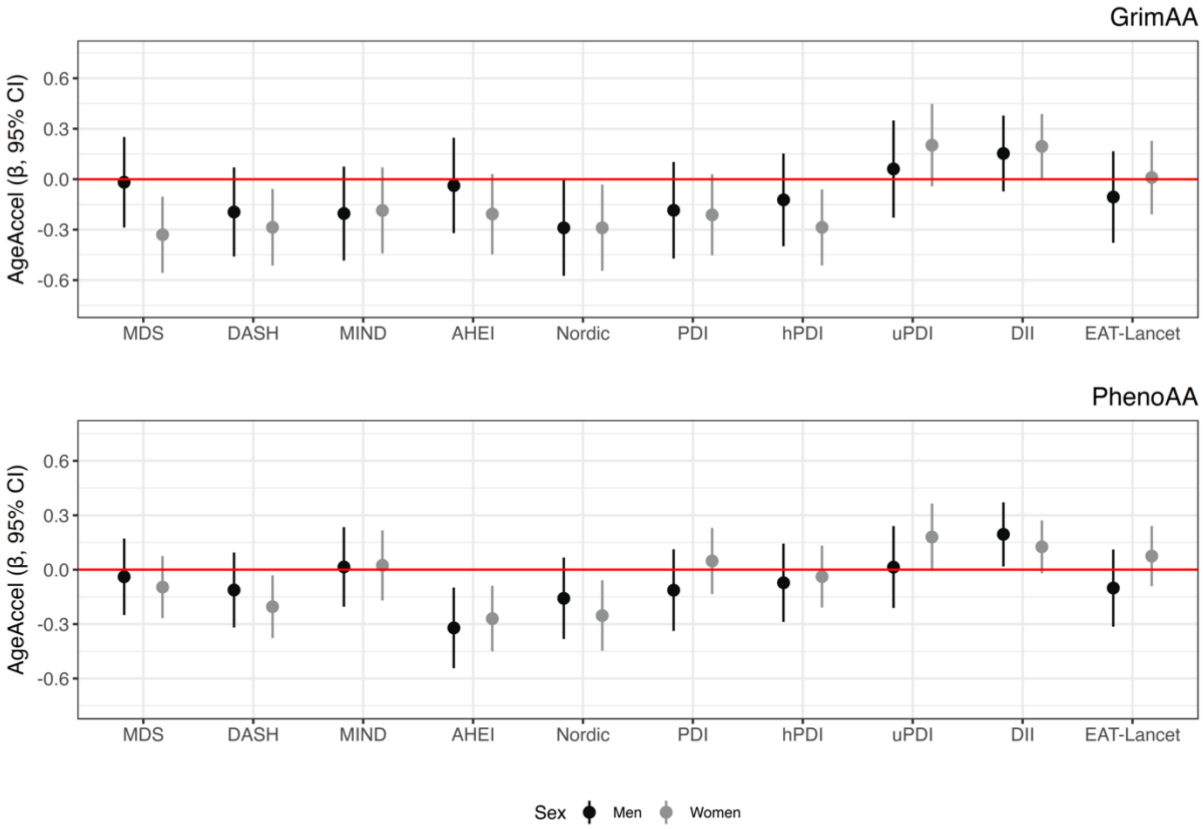

***Supplementary Figure 4: Associations between recommendation-based diet quality scores and the two measures of epigenetic age acceleration stratified by sex.***

Plots display the β-coefficients and 95% CIs from adjusted linear regression models, which represent the adjusted mean difference (in years) for the epigenetic acceleration metrics per 1-SD increase in diet quality among women (grey lines; n= 3665) and men (black lines; n= 2805). Models was adjusted for batch effect, blood cell proportion, smoking status and total energy intake. Models testing associations with the DASH, Nordic, EAT-Lancet, PDI, hPDI and uPDI scores were additionally adjusted for alcohol intake (g/day). Significant statistical interaction determined by cross-product terms were observed for sex on the relationship between the DII and GrimAA (p-interaction= 0.04/ FDR-adjusted p-interaction= 0.82).

Abbreviation: GrimAA, DNA methylation GrimAge Acceleration; PhenoAA, DNA methylation PhenoAge Acceleration; MDS, Mediterranean Diet Score; DASH, Dietary Approaches to Stop Hypertension; MIND, Mediterranean-DASH Intervention for Neurodegenerative Delay; AHEI, Alternate Healthy Eating Index score; PDI, Plant-based Diet Index; hPDI, Healthful Plant-based Diet Index; uPDI, Unhealthful Plant-based Diet Index; DII, Dietary Inflammatory Index.

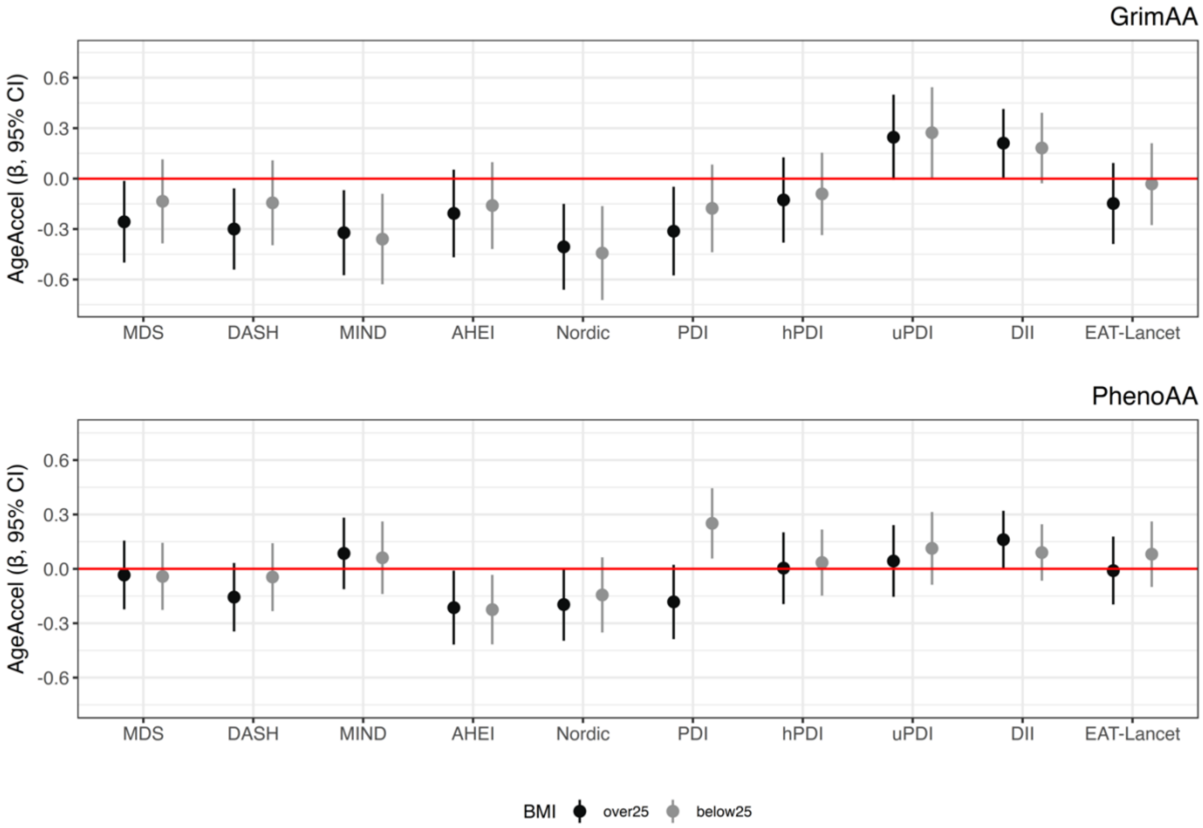

***Supplementary Figure 5: Associations between recommendation-based diet quality scores and the two measures of epigenetic age acceleration stratified by body mass index.***

Plots display the β-coefficients and 95% CIs from adjusted linear regression models, which represent the adjusted mean difference (in years) for the epigenetic acceleration metrics per 1-SD increase in diet quality among participants who have BMIs < 25 (grey lines; n= 3037) and ≥25 (black lines; n= 3398). Thirty-five participants were missing information on body mass index. Models was adjusted for batch effect, blood cell proportion, sex, smoking status and total energy intake. Models testing associations with the DASH, Nordic, EAT-Lancet, PDI, hPDI and uPDI scores were additionally adjusted for alcohol intake (g/day). Significant statistical interaction determined by cross-product terms were observed for body mass index on the relationship between the PDI and PhenoAA (p-interaction= 0.004/ FDR-adjusted p-interaction= 0.08).

Abbreviation: GrimAA, DNA methylation GrimAge Acceleration; PhenoAA, DNA methylation PhenoAge Acceleration; MDS, Mediterranean Diet Score; DASH, Dietary Approaches to Stop Hypertension; MIND, Mediterranean-DASH Intervention for Neurodegenerative Delay; AHEI, Alternate Healthy Eating Index score; PDI, Plant-based Diet Index; hPDI, Healthful Plant-based Diet Index; uPDI, Unhealthful Plant-based Diet Index; DII, Dietary Inflammatory Index.

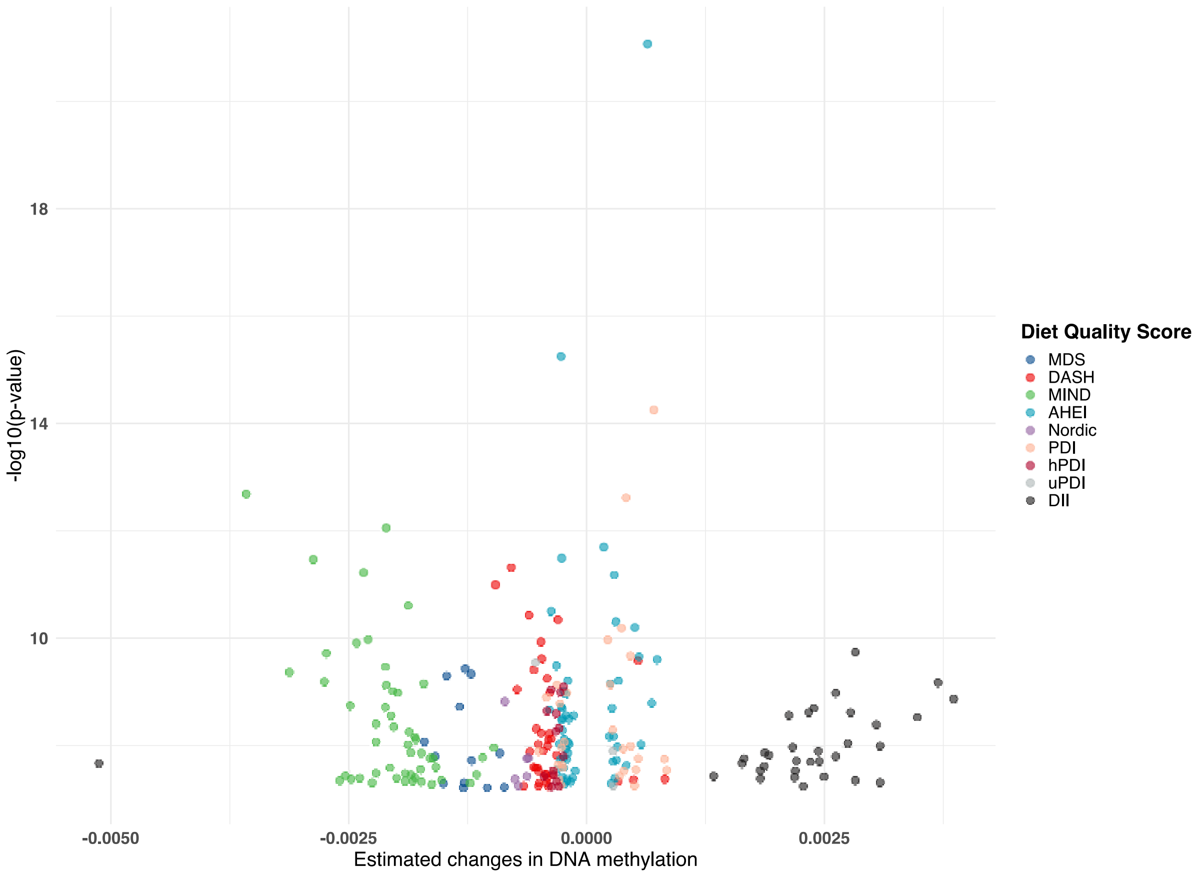

***Supplementary Figure 6: Methylation beta estimates values across the recommendation-based diet quality scores.***

Volcano plot depicting beta coefficients and -log10(p values) of CpGs demonstrating epigenome-wide significance, determined by a Bonferroni threshold (5.95e-08, corrected for multiple testing at 0.05 divided by the number of CpGs), across all diet quality scores. Linear models were adjusted for age, sex, batch effects, blood cell proportion, the first ten genetic principal components (to account for population stratification), and smoking status. Negative coefficient estimates indicate decreased methylation, while positive estimates designate increased methylation.

Abbreviation: MDS, Mediterranean Diet Score; DASH, Dietary Approaches to Stop Hypertension; MIND, Mediterranean-DASH Intervention for Neurodegenerative Delay; AHEI, Alternate Healthy Eating Index score; PDI, Plant-based Diet Index; hPDI, Healthful Plant-based Diet Index; uPDI, Unhealthful Plant-based Diet Index; DII, Dietary Inflammatory Index

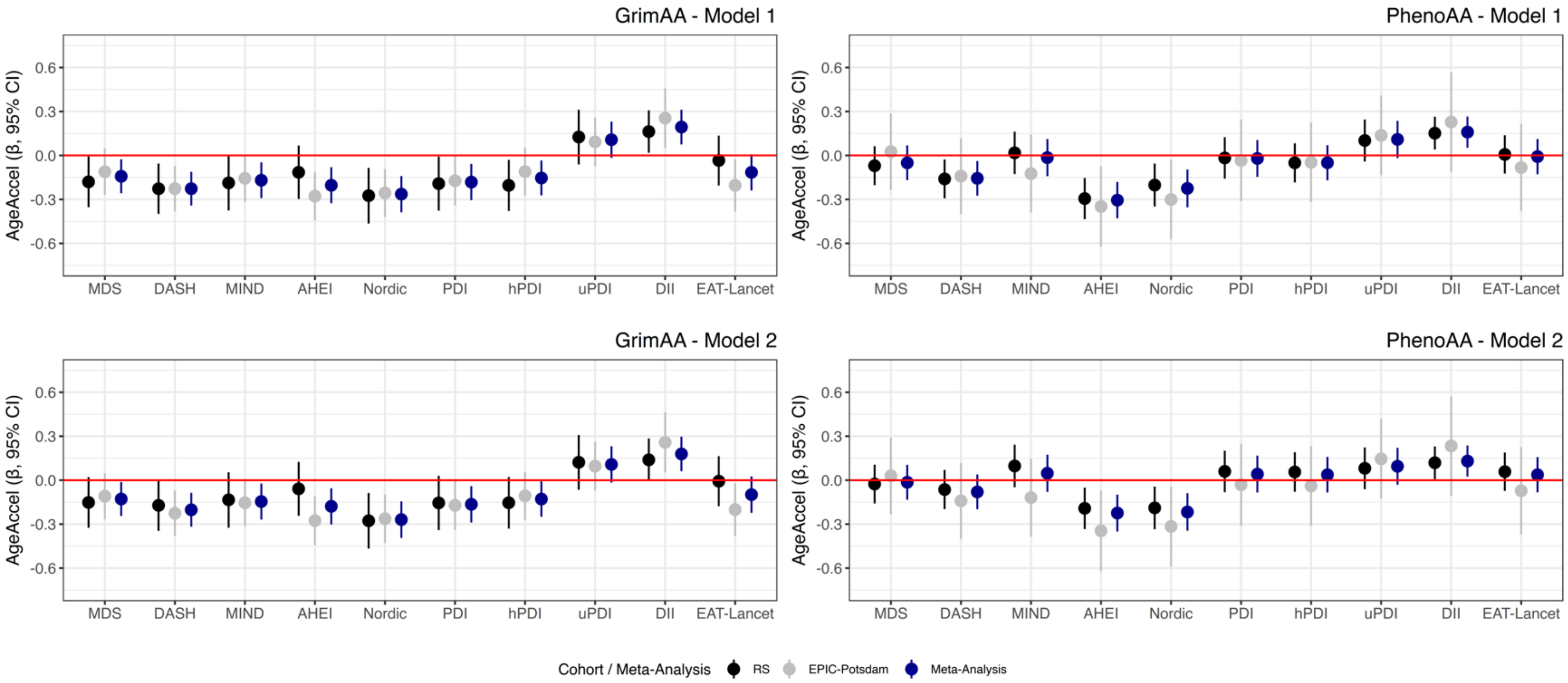

***Supplementary Figure 7: Associations between recommendation-based diet quality scores and the two measures of epigenetic age acceleration across discovery and replication cohorts and meta-analyses results.***

Plots display the β-coefficients and 95% CIs from adjusted linear regression models, which represent the adjusted mean difference (in years) for the epigenetic acceleration metrics per 1-SD increase in diet quality among participants from RS (black lines; n= 3665) and EPIC-Potsdam (grey lines; n= 1034), and pooled meta-analysis results from both cohorts (blue lines). Model 1 was adjusted for batch effect, blood cell proportion, sex, smoking status and total energy intake. Models testing associations with the DASH, Nordic, EAT-Lancet, PDI, hPDI and uPDI scores were additionally adjusted for alcohol intake (g/day). Model 2 was additionally adjusted for body mass index. The meta-analysis was conducted using an inverse-variance weighted approach to estimate pooled associations across the two cohorts.

Abbreviation: MDS, Mediterranean Diet Score; DASH, Dietary Approaches to Stop Hypertension; MIND, Mediterranean-DASH Intervention for Neurodegenerative Delay; AHEI, Alternate Healthy Eating Index score; PDI, Plant-based Diet Index; hPDI, Healthful Plant-based Diet Index; uPDI, Unhealthful Plant-based Diet Index; DII, Dietary Inflammatory Index; RS, Rhineland Study.

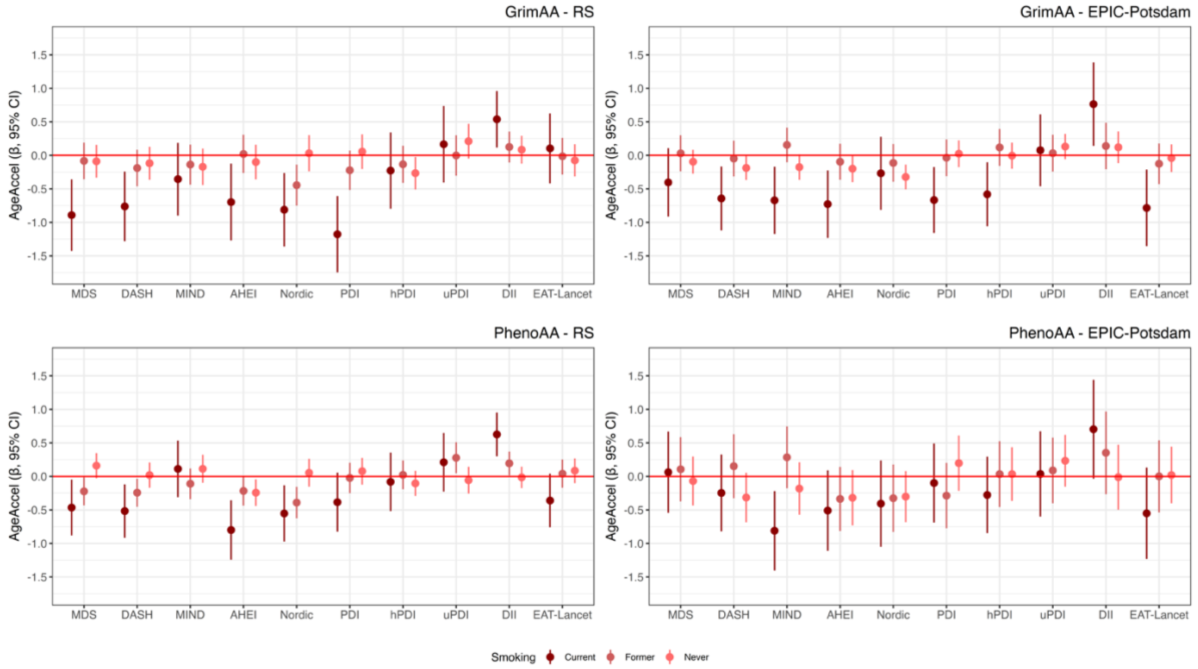

***Supplementary Figure 8: Associations between recommendation-based diet quality scores and the two measures of epigenetic age acceleration stratified by smoking status across discovery and replication cohorts.*** Plots display the β-coefficients and 95% CIs from adjusted linear regression models, which represent the adjusted mean difference (in years) for the epigenetic acceleration metrics per 1-SD increase in diet quality among current smokers (dark red lines; RS n= 767, EPIC-Potsdam n=210), former smokers (red lines; RS n= 2653,  EPIC-Potsdam n=315), and never smokers (light red lines; RS n= 3050, EPIC-Potsdam n=509). Models were adjusted for batch effect, blood cell proportion, sex and total energy intake. Models testing associations with the DASH, Nordic, EAT-Lancet, PDI, hPDI and uPDI scores were additionally adjusted for alcohol intake (g/day).

Abbreviation: MDS, Mediterranean Diet Score; DASH, Dietary Approaches to Stop Hypertension; MIND, Mediterranean-DASH Intervention for Neurodegenerative Delay; AHEI, Alternate Healthy Eating Index score; PDI, Plant-based Diet Index; hPDI, Healthful Plant-based Diet Index; uPDI, Unhealthful Plant-based Diet Index; DII, Dietary Inflammatory Index; RS, Rhineland Study

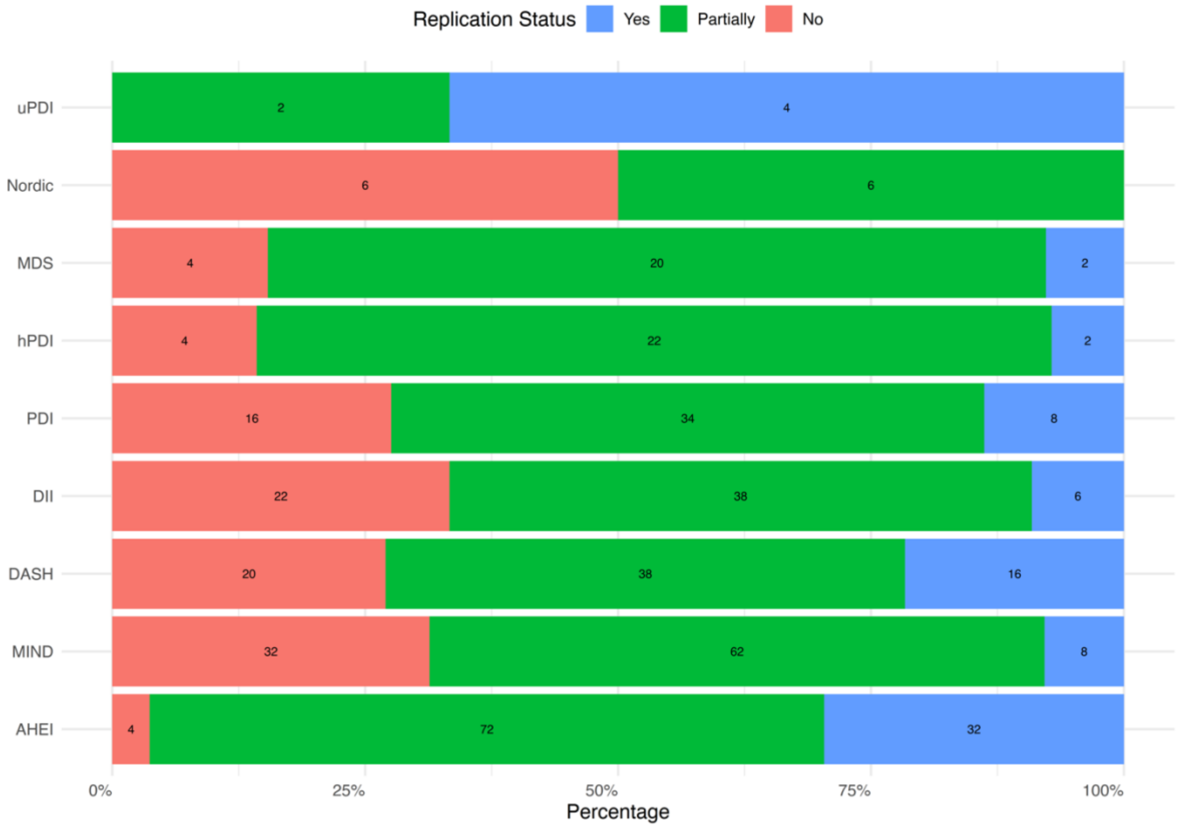

***Supplementary Figure 9: Replication rates of epigenome-wide significant CpGs in the EPIC-Potsdam cohort, stratified by dietary pattern.***

This stacked bar plot displays the percentage of epigenome-wide significant CpGs identified in the Rhineland Study that were successfully replicated in EPIC-Potsdam ($p < 0.05$ with consistent effect direction), categorized by diet quality score. CpGs were classified into three replication categories: "Yes" (blue) for CpGs meeting the replication criteria, "Partially" (green) for CpGs with consistent effect directions but $p > 0.05$ in the replication cohort, and "No" (red) for CpGs that did not replicate. The numbers within the bars indicate the absolute count of CpGs within each replication category for each dietary pattern.

Abbreviation: MDS, Mediterranean Diet Score; DASH, Dietary Approaches to Stop Hypertension; MIND, Mediterranean-DASH Intervention for Neurodegenerative Delay; AHEI, Alternate Healthy Eating Index score; PDI, Plant-based Diet Index; hPDI, Healthful Plant-based Diet Index; uPDI, Unhealthful Plant-based Diet Index; DII, Dietary Inflammatory Index.

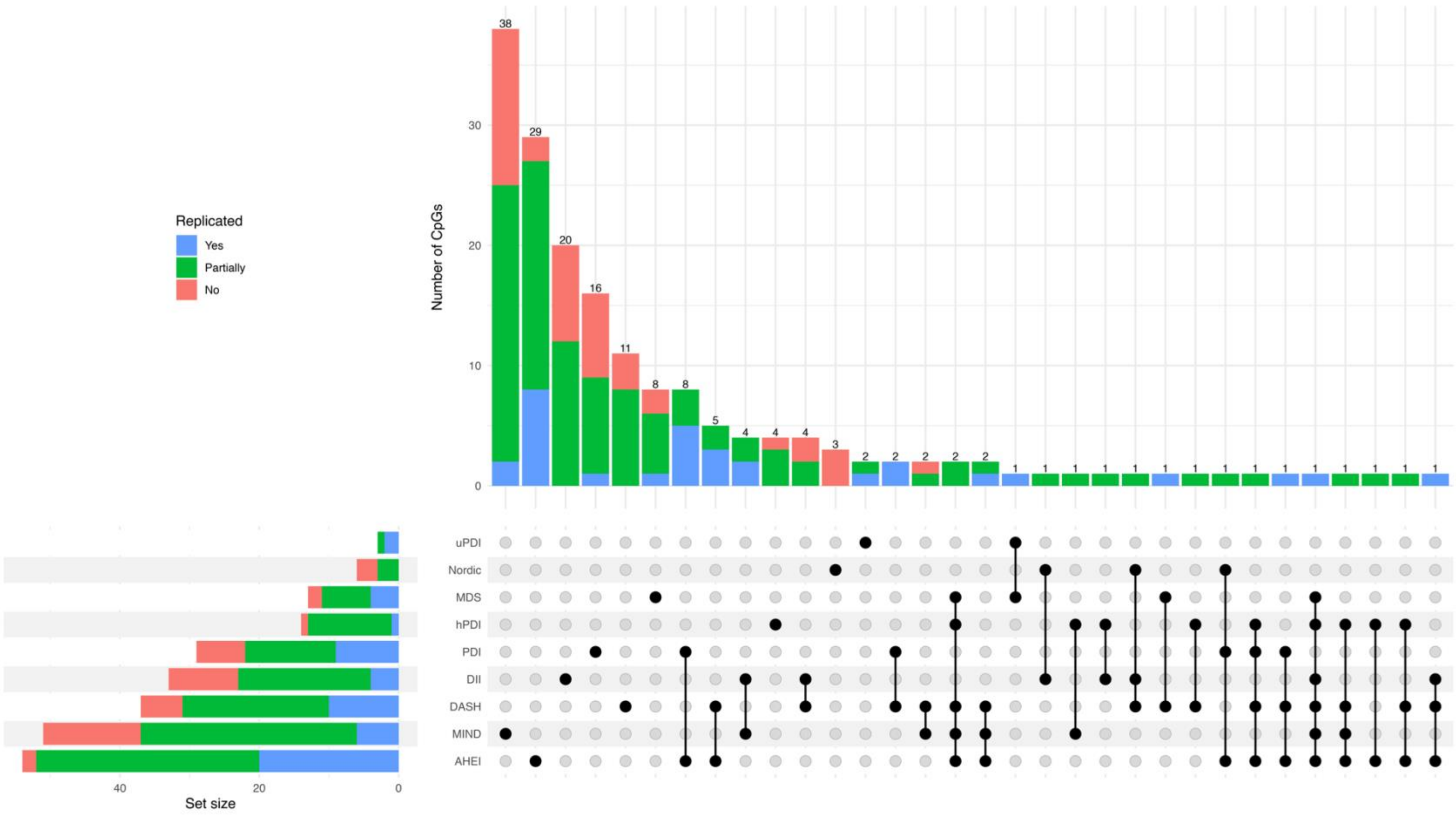

***Supplementary Figure 10: Overlap and replication status of diet-associated DNA methylation sites (epigenome-wide significant CpGs) across recommendation-based diet quality scores.***

The UpSet plot summarizes the overlap of differentially methylated CpGs identified in the discovery cohort across the recommendation-based diet quality scores while incorporating their replication status in the EPIC-Potsdam cohort. The bottom left horizontal bar graph displays the total number of epigenome-wide significant CpGs identified for each diet quality score. The matrix panel at the bottom right represents CpGs associated with specific diet quality scores, where dots indicate unique CpGs and connected dots highlight shared CpGs across multiple dietary patterns. The top bar graph recapitulates the number of differentially methylated CpGs for each unique or overlapping combination. The bars in the top graph and bottom left graph are color-coded to represent replication status in the EPIC-Potsdam cohort: "Yes" (blue) for CpGs meeting the replication criteria, "Partially" (green) for CpGs with consistent effect directions but $p > 0.05$ in the replication cohort, and "No" (red) for CpGs that did not replicate.

If a CpG site was associated with multiple diet quality scores but exhibited different replication statuses across diets, its replication status was determined by the highest level of replication observed.

Abbreviations: MDS, Mediterranean Diet Score; DASH, Dietary Approaches to Stop Hypertension; MIND, Mediterranean-DASH Intervention for Neurodegenerative Delay; AHEI, Alternate Healthy Eating Index; PDI, Plant-based Diet Index; hPDI, Healthful Plant-based Diet Index; uPDI, Unhealthful Plant-based Diet Index; DII, Dietary Inflammatory Index.

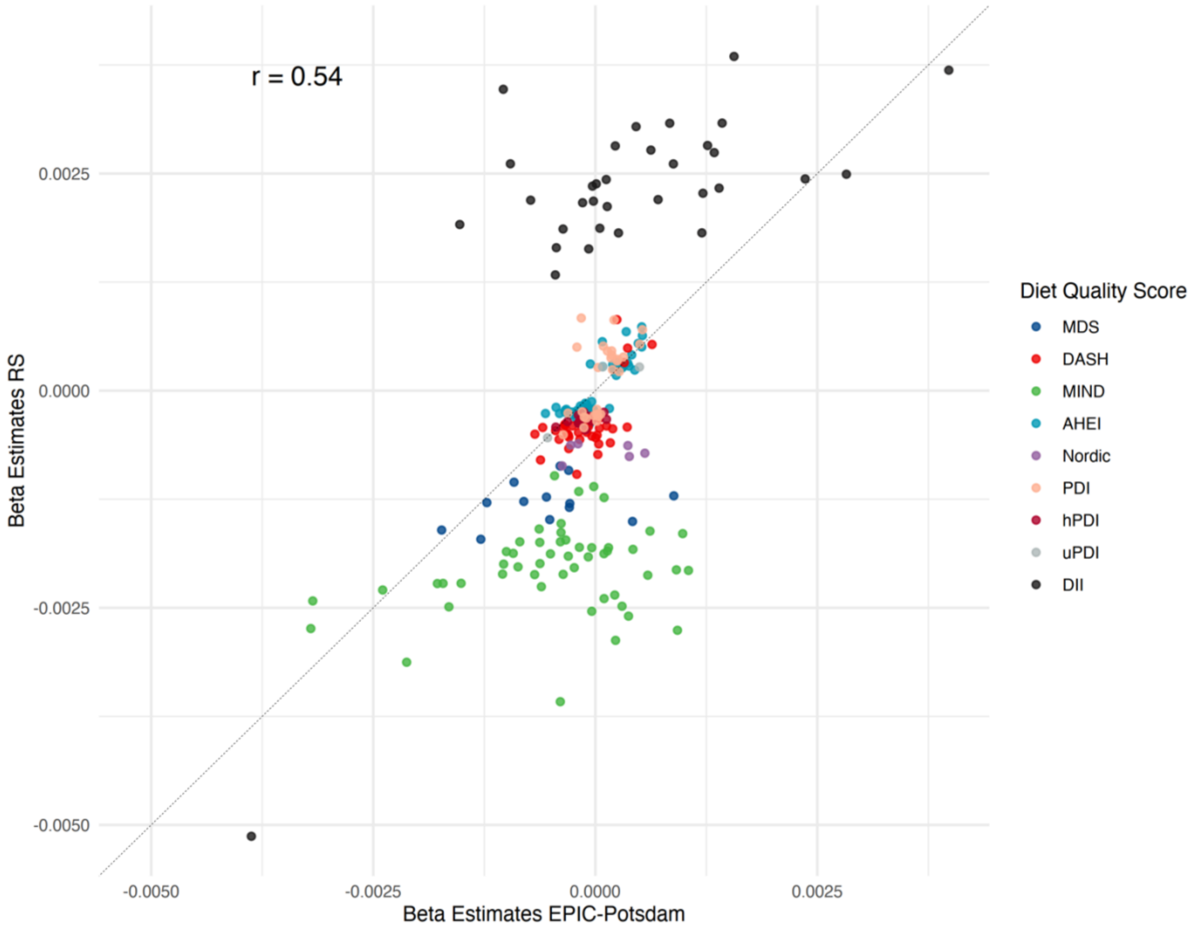

***Supplementary Figure 11: Beta-beta correlation of epigenome-wide significant CpGs between the Rhineland Study and EPIC-Potsdam cohorts.***

The scatter plot visualizes the correlation of beta estimates for CpG sites that reached epigenome-wide significance ($p < 5.95 \times 10^{-8}$) in the discovery cohort (Rhineland Study) with their corresponding beta estimates in the replication cohort (EPIC-Potsdam). Each point represents a CpG site, color-coded according to its association with a specific diet quality score. The diagonal dashed line represents the line of identity (y = x), indicating perfect concordance of effect estimates between cohorts. The Pearson correlation coefficient (r = 0.54) quantifies the overall correlation between effect estimates across studies.

Abbreviations: MDS, Mediterranean Diet Score; DASH, Dietary Approaches to Stop Hypertension; MIND, Mediterranean-DASH Intervention for Neurodegenerative Delay; AHEI, Alternate Healthy Eating Index; PDI, Plant-based Diet Index; hPDI, Healthful Plant-based Diet Index; uPDI, Unhealthful Plant-based Diet Index; DII, Dietary Inflammatory Index; RS, Rhineland Study.

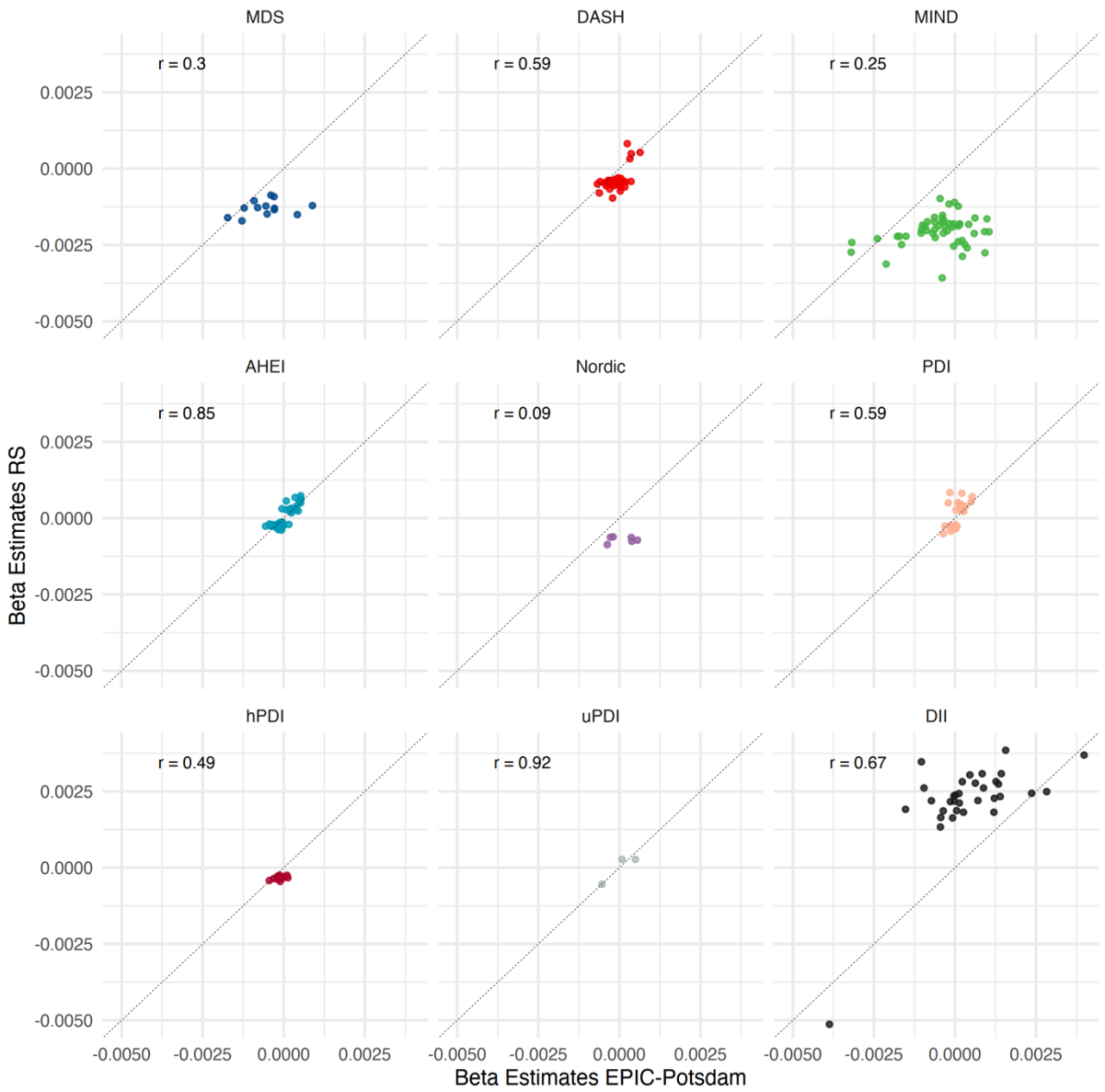

***Supplementary Figure 12: Beta-beta correlation of epigenome-wide significant CpGs between the Rhineland Study and EPIC-Potsdam cohorts, stratified by diet quality score.***

Each scatter plot presents the correlation of beta estimates for CpG sites that reached epigenome-wide significance ($p < 5.95 \times 10^{-8}$) in the discovery cohort (Rhineland Study) with their corresponding beta estimates in the replication cohort (EPIC-Potsdam), stratified by diet quality score. Each point represents a CpG site, and the diagonal dashed line ($y = x$) represents perfect concordance of effect estimates between cohorts. Pearson correlation coefficients ($r$) are displayed for each diet quality score to quantify the strength of association across studies.

Abbreviations: MDS, Mediterranean Diet Score; DASH, Dietary Approaches to Stop Hypertension; MIND, Mediterranean-DASH Intervention for Neurodegenerative Delay; AHEI, Alternate Healthy Eating Index; PDI, Plant-based Diet Index; hPDI, Healthful Plant-based Diet Index; uPDI, Unhealthful Plant-based Diet Index; DII, Dietary Inflammatory Index; RS, Rhineland Study

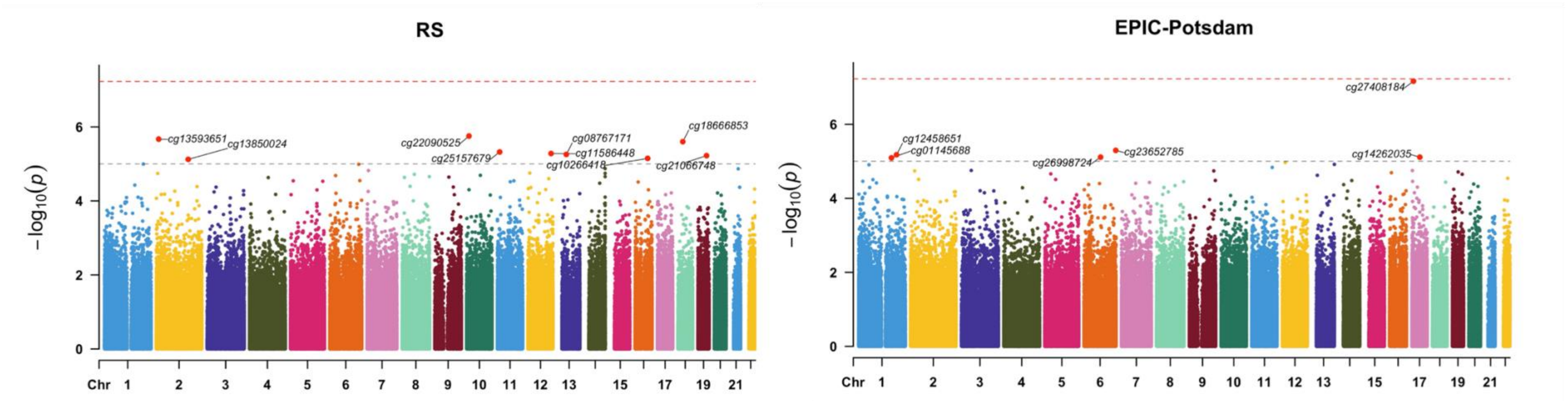

***Supplementary Figure 13: Supplementary Figure 17: Manhattan plots of the epigenome-wide associations for the EAT-Lancet diet score in the Rhineland Study and EPIC-Potsdam cohorts.***

Manhattan plots display the genome-wide associations between the EAT-Lancet diet score and DNA methylation levels in the Rhineland Study (left) and EPIC-Potsdam cohort (right). The y-axis represents the negative log-transformed p-values, while the x-axis shows the genomic position of CpG sites. The red horizontal line denotes the epigenome-wide significance threshold based on Bonferroni correction ($p < 5.95 \times 10^{-8}$), calculated as 0.05 divided by the number of tested CpGs. The black horizontal line denotes the suggestive significance threshold ($p < 1 \times 10^{-5}$). Linear models were adjusted for age, sex, batch effects, blood cell proportions, the first ten genetic principal components to account for population stratification, and smoking status. The top CpG sites on each chromosome are annotated.

Abbreviations: RS, Rhineland Study.

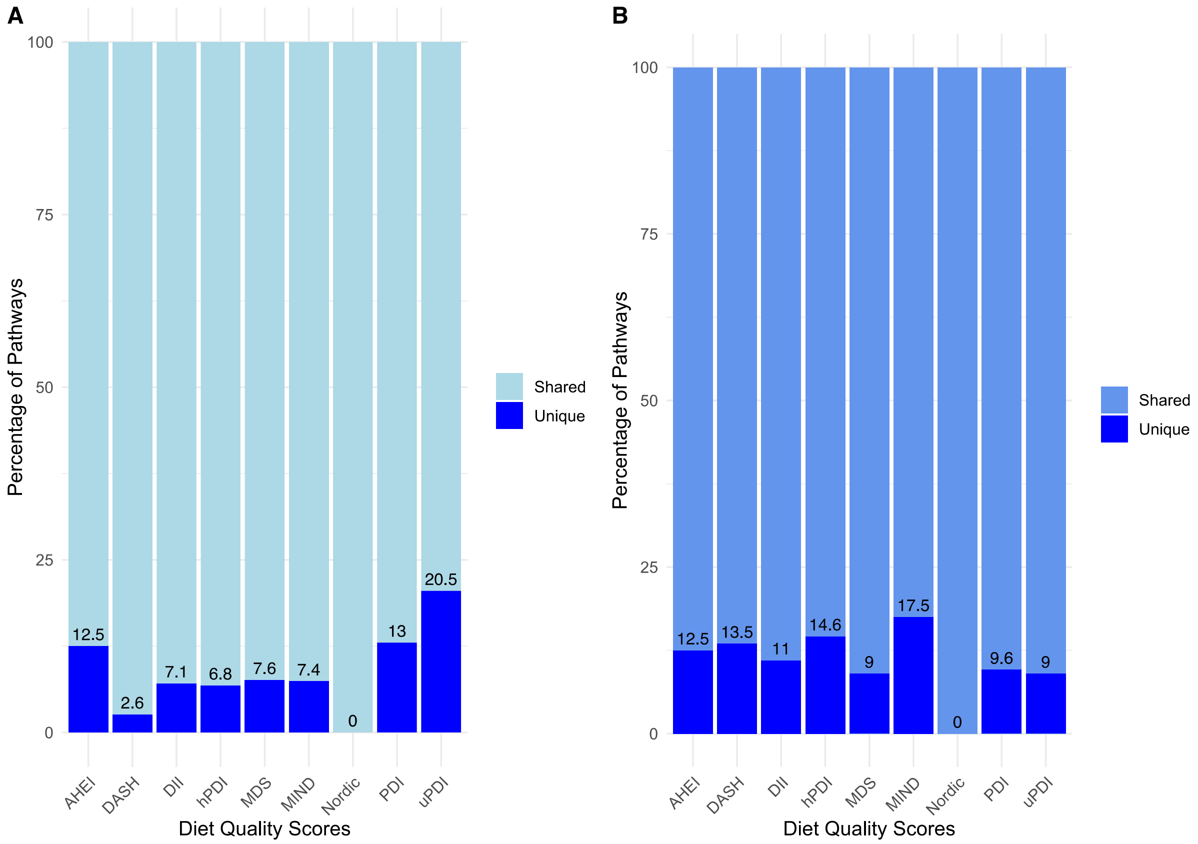

***Supplementary Figure 14: Percentage of unique and shared pathways associated with recommendation-based diet quality scores identified through by KEGG and GO databases.***

Bar plots for A) KEGG and B) GO pathways.

Abbreviation: MDS, Mediterranean Diet Score; DASH, Dietary Approaches to Stop Hypertension; MIND, Mediterranean-DASH Intervention for Neurodegenerative Delay; AHEI, Alternate Healthy Eating Index score; PDI, Plant-based Diet Index; hPDI, Healthful Plant-based Diet Index; uPDI, Unhealthful Plant-based Diet Index; DII, Dietary Inflammatory Index; KEGG, Kyoto Encyclopedia of Genes and Genomes; GO, Gene Ontology.

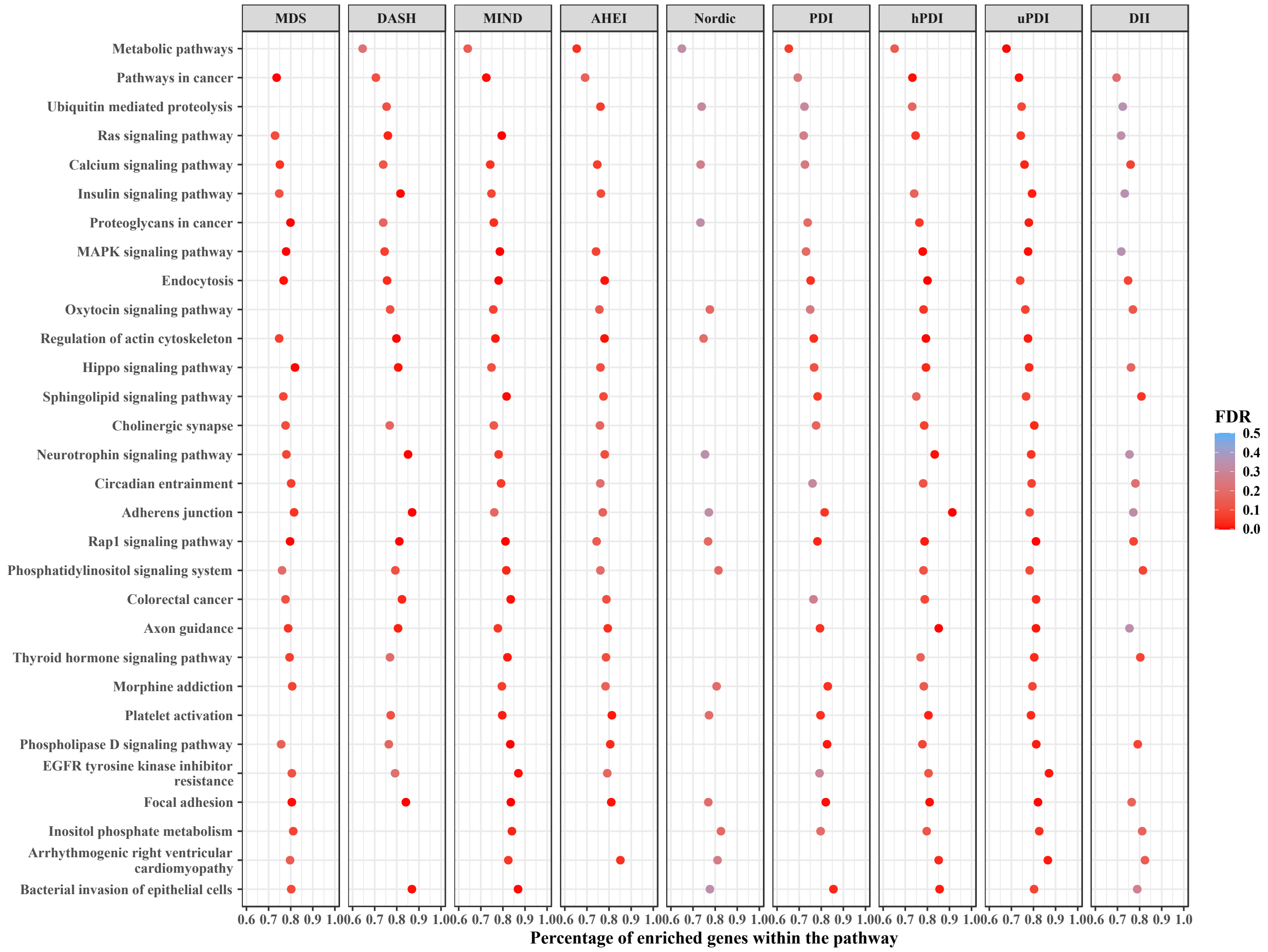

***Supplementary Figure 15: Shared KEGG pathways identified in the enrichment analyses across at least seven recommendation-based diet quality scores.***

Abbreviation: MDS, Mediterranean Diet Score; DASH, Dietary Approaches to Stop Hypertension; MIND, Mediterranean-DASH Intervention for Neurodegenerative Delay; AHEI, Alternate Healthy Eating Index score; PDI, Plant-based Diet Index; hPDI, Healthful Plant-based Diet Index; uPDI, Unhealthful Plant-based Diet Index; DII, Dietary Inflammatory Index; KEGG, Kyoto Encyclopedia of Genes and Genomes.

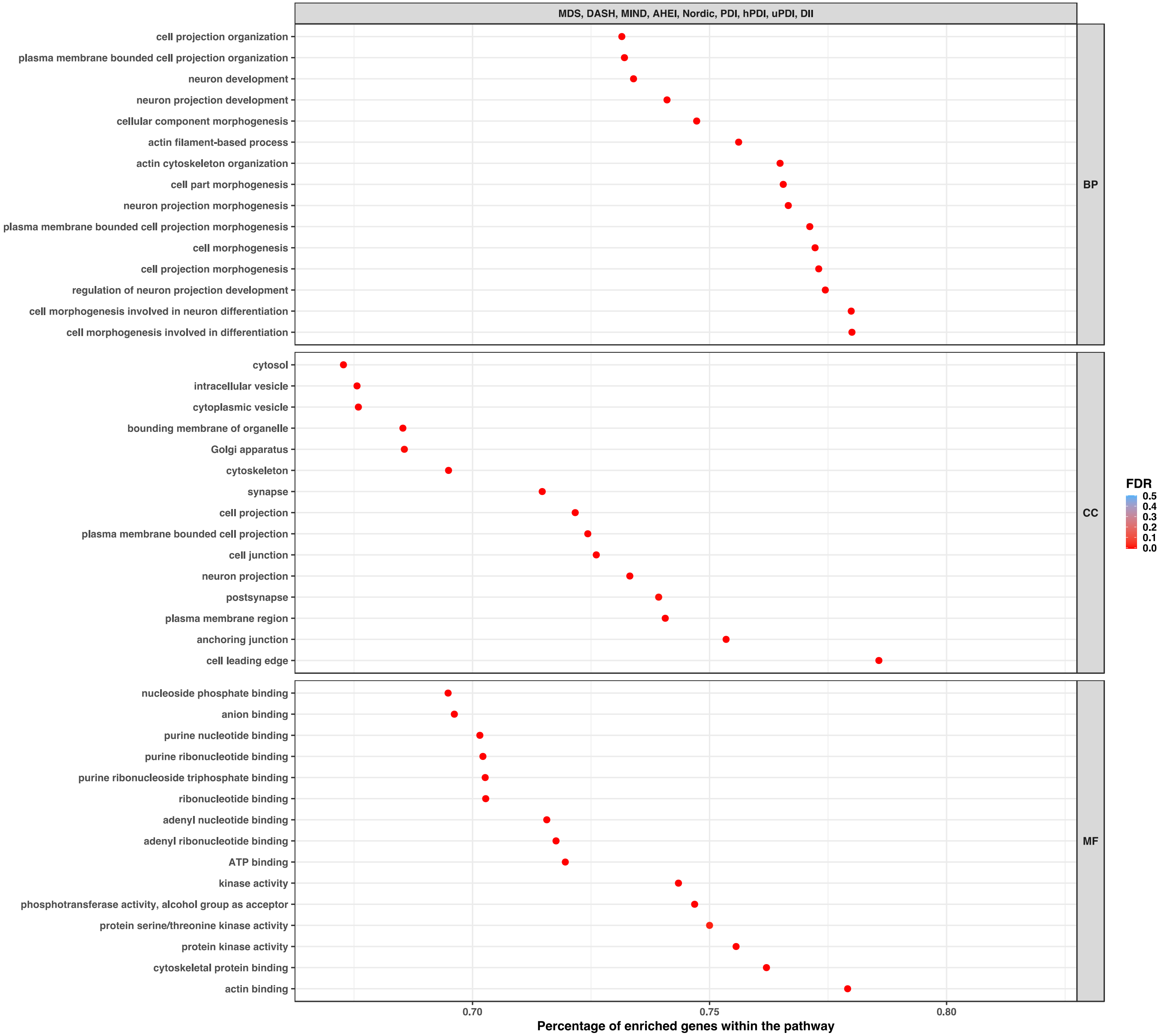
MDS, DASH, MIND, AHEI, Nordic, PDI, hPDI, uPDI, DII
cell projection organization
plasma membrane bounded cell projection organization
neuron development
neuron projection development
cellular component morphogenesis
actin filament-based process
actin cytoskeleton organization
cell part morphogenesis
neuron projection morphogenesis
plasma membrane bounded cell projection morphogenesis
cell morphogenesis
cell projection morphogenesis
regulation of neuron projection development
cell morphogenesis involved in neuron differentiation
cell morphogenesis involved in differentiation
BP
cytosol
intracellular vesicle
cytoplasmic vesicle
bounding membrane of organelle
Golgi apparatus
cytoskeleton
synapse
cell projection
plasma membrane bounded cell projection
cell junction
neuron projection
postsynapse
plasma membrane region
anchoring junction
cell leading edge
CC
nucleoside phosphate binding
anion binding
purine nucleotide binding
purine ribonucleotide binding
purine ribonucleoside triphosphate binding
ribonucleotide binding
adenyl nucleotide binding
adenyl ribonucleotide binding
ATP binding
kinase activity
phosphotransferase activity, alcohol group as acceptor
protein serine/threonine kinase activity
protein kinase activity
cytoskeletal protein binding
actin binding
MF
0.70
0.75
0.80
Percentage of enriched genes within the pathway
FDR
0.5
0.4
0.3
0.2
0.1
0.0

***Supplementary Figure 16: Shared GO pathways identified in the enrichment analyses across all recommendation-based diet quality scores, according to distinct ontologies.*** Abbreviation: MDS, Mediterranean Diet Score; DASH, Dietary Approaches to Stop Hypertension; MIND, Mediterranean-DASH Intervention for Neurodegenerative Delay; AHEI, Alternate Healthy Eating Index score; PDI, Plant-based Diet Index; hPDI, Healthful Plant-based Diet Index; uPDI, Unhealthful Plant-based Diet Index; DII, Dietary Inflammatory Index; GO, Gene Ontology; BP, Biological Process; CC, Cellular Component; MF Molecular Function.

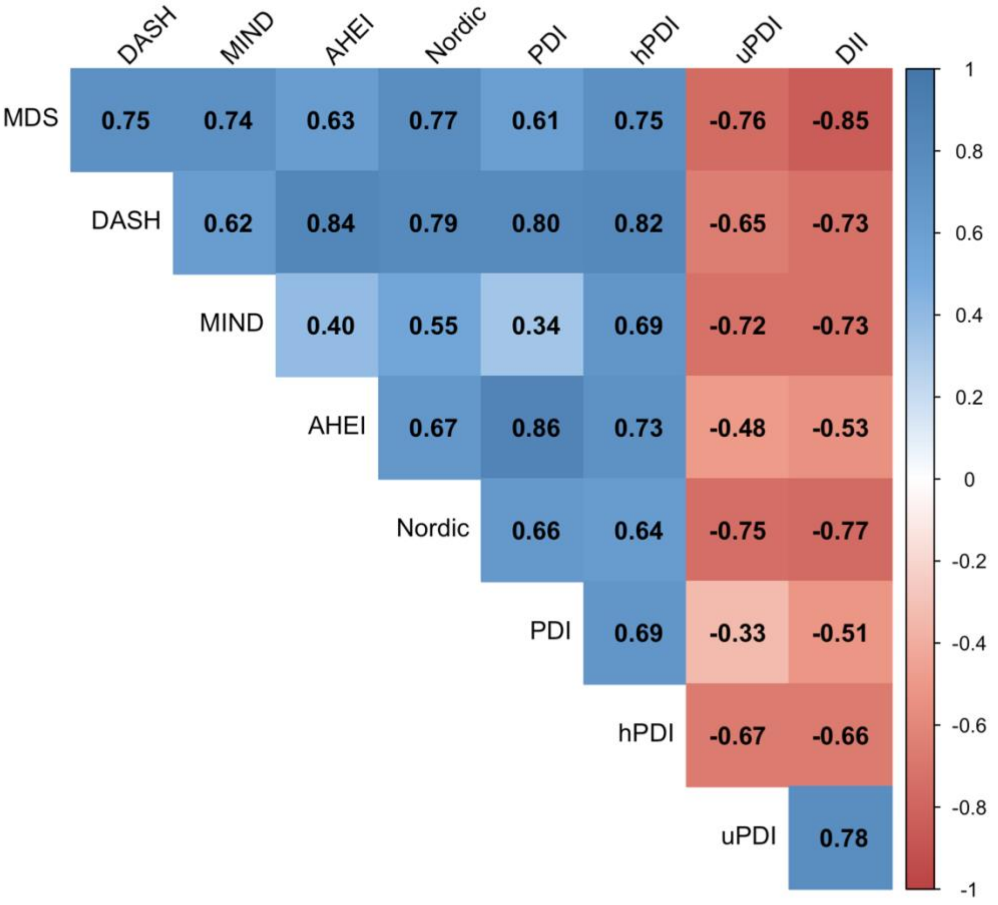

***Supplementary Figure 17: Pairwise Pearson correlations of beta coefficients for epigenome-wide significant CpGs across all diet quality scores.***

The heatmap displays the strength and direction of correlations between the effect estimates (beta values) of diet-associated CpGs with epigenome-wide significance ($p < 5.95 \times 10^{-8}$). Each cell shows the Pearson correlation coefficient (r) between a pair of diets, calculated across all CpGs that reached significance in at least one diet. Red colors indicate negative correlations, while blue indicate positive correlations, and the intensity reflects the strength of the association.

Abbreviation: MDS, Mediterranean Diet Score; DASH, Dietary Approaches to Stop Hypertension; MIND, Mediterranean-DASH Intervention for Neurodegenerative Delay; AHEI, Alternate Healthy Eating Index score; PDI, Plant-based Diet Index; hPDI, Healthful Plant-based Diet Index; uPDI, Unhealthful Plant-based Diet Index; DII, Dietary Inflammatory Index.

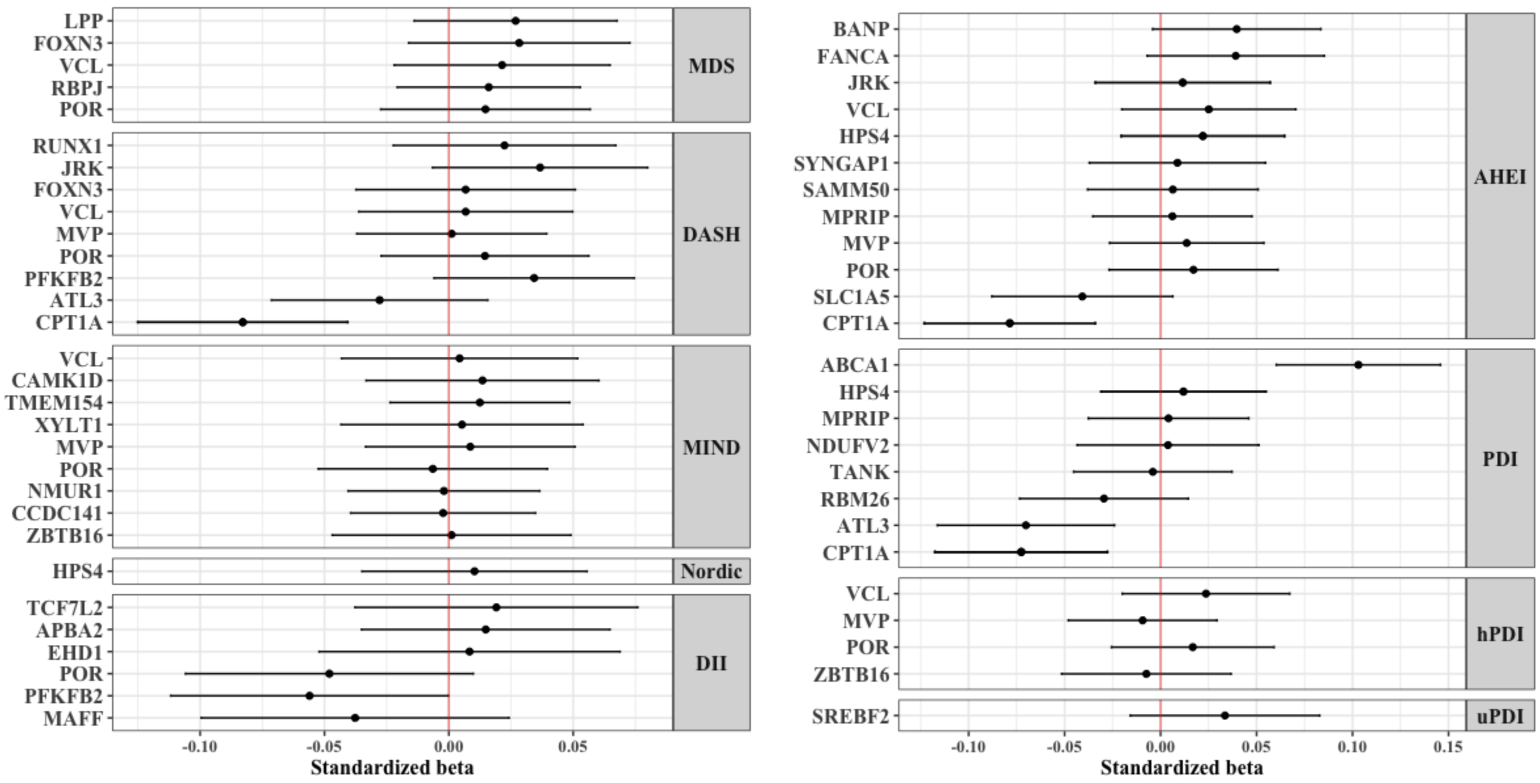

***Supplementary Figure 18: The association between diet quality scores and the corresponding gene expression levels***

Forest plots showing the effect estimate of genes on diet quality scores obtained through the following multivariable linear model: $Gene_y$ ~ intercept + Diet quality $score_x$ + age + sex + total energy intake + blood cell proportion + batch effect. Abbreviation: MDS, Mediterranean Diet Score; DASH, Dietary Approaches to Stop Hypertension; MIND, Mediterranean-DASH Intervention for Neurodegenerative Delay; AHEI, Alternate Healthy Eating Index score; PDI, Plant-based Diet Index; hPDI, Healthful Plant-based Diet Index; uPDI, Unhealthful Plant-based Diet Index; DII, Dietary Inflammatory Index.

### Supplementary Methods

#### Dietary Assessment

Habitual dietary intake was assessed by a self-administered semi-quantitative food frequency questionnaire (FFQ). The questionnaire was originally developed for the European Prospective Investigation into Cancer and Nutrition (EPIC) study in Potsdam and adapted for the Rhineland Study[1]. Participants were questioned about their habitual intake of 132 food and beverage items in the last 12 months, including standard portion sizes. Depending on the food item, there were between four and eleven frequency options available, ranging from "never" to "11 times per day or more". Additionally, participants were also asked about fat contents of consumed dairy and meat products, as well as types of fat used for food preparation.

The FFQs were eligible for inclusion in analyses provided that information was available for at least 80% of core food items. Fats used for food preparation or additives to hot beverages were not considered as core food items. Missing data on the FFQ were found for 118 core food items, with a maximum of 31 missing values in one FFQ item. Most food items had only 1 or 2 missing values. To retain all observations with ≥80% of questionnaire competition, missing values on the FFQ were imputed using MissForest.[2] Using SAS 9.4, we computed the sum intake of each food/beverage and estimated macro- and micronutrients, water, and energy intake based on data from the German Food Code and Nutrient Database (version 3.02).[3] The dietary data were categorized into 17 main food groups and 71 subgroups in accordance with the EPICSOFT classification scheme.[4]

#### Diet Quality Scores

The MDS incorporates foods that have demonstrated a protective effect against cardiovascular and other chronic diseases.[5] It is based on nine key components. A score of 1 point was assigned to individuals whose intake of vegetables, fruits, nuts, legumes, whole grains, fish, and the MUFA to SFA ratio exceeded the median. Similarly, a score of 1 point was given to those whose intake of red and processed meats, as well as dairy, fell below the median. For alcohol, a value of 1 was assigned to men consuming between 10 and 50 grams per day and women consuming between 5 and 25 grams per day. The cumulative points from all components were used to calculate the MDS score, which ranged from 0 to 9, with higher scores indicating healthier dietary patterns.

The DASH diet incorporates various foods and nutrients known to protect against hypertension.[6] For the components of fruits, vegetables, whole grains, nuts and legumes, and low-fat dairy, individuals in the lowest quintile of consumption received 1 point, with an additional point awarded for each increasing quintile. Conversely, individuals in the highest quintile of consumption for red and processed meats, sugar-sweetened beverages, and sodium were given 1 point, with an additional point added for each decreasing quintile. The DASH diet component scores were summed, resulting in a potential score range of 8 to 40.

The MIND diet is a dietary approach that combines elements of the Mediterranean diet and the DASH diet, focusing on specific foods believed to support brain health and reduce the risk of cognitive decline.[7] The score contains recommendations regarding 15 food components, including 10 food components considered to be healthy for the brain (i.e., green leafy vegetables, other vegetables, nuts, berries, beans, whole grains, fish, poultry, olive oil, and wine) and five unhealthy food components (i.e., red meat, butter and stick margarine, cheese, fast fried food, and pastries and sweets). Since we did not have specific data on olive oil consumption, we did not consider it for calculating the score. For each other food component, a 0 was assigned if participants did not adhere to the recommendations, a 0.5 for moderate adherence, and a 1 for good adherence. Scores assigned to each food component were summed, obtaining a total score ranging from 0 to 14.

The AHEI score incorporates additional evidence regarding the relationship between diet and chronic disease in order to enhance its predictive ability.[8] This scoring system is based on the intake levels of 11 specific foods and nutrients. Higher scores were assigned to individuals with greater consumption of vegetables, fruits, whole grains, nuts and legumes, PUFAs, and omega-3 fatty acids. Conversely, lower scores were given to individuals with higher intake of sugar-sweetened beverages, red and processed meats, trans fatty acids, and sodium. Moderate alcohol intake was considered favourable and awarded a moderate score. However, in the present study constructed a modified score by excluding the trans-fat component because it was unavailable in the Rhineland Study dietary data. Each component received a score ranging from 0 (indicating the least favourable) to 10 (indicating the most favourable), with partial scores assigned proportionally based on intake levels. The scores for each component were then added together to calculate the total AHEI score which has a potential range of 0 to 100.

The Nordic diet tries to reflect the eating pattern followed in Nordic countries that emphasizes consuming locally sourced, seasonal, and nutrient-rich foods.[9] The score ranges from 0 to 18 points, incorporating 9 components: whole grain and rye bread, berries, apples and pears, fish, cabbage and cruciferous vegetables, root vegetables, low-fat dairy products, potatoes, and vegetable fats. Each food component is categorized into sex-specific tertiles of intake and the participants then received a score of 0 to 2 points according to the first, second, and third tertile, respectively.

The EAT-Lancet diet was proposed in 2019 by the EAT-Lancet Commission on Food, Planet, and Health. The Commission articulated global scientific goals geared towards cultivating both healthy dietary practices and sustainable food systems. These objectives delineated a secure zone within which dietary consumption could operate while concurrently considering human and planetary health. The EAT-Lancet diet score was calculated similarly to Knuppel et al, based on the EAT-Lancet Commission recommendations.[10,11] This score is comprised of 14 dietary components, encompassing grains, tubers and starchy vegetables, vegetables, fruits, dairy products, red meat, poultry, eggs, fish, legumes, soy foods, nuts, fats, and sugar/sugary products. To ensure consistency with the EAT-Lancet Commission's recommendations, the intake for each dietary component was standardized to 2500 kcal. Participants were awarded 1 point for adhering to each of these recommendations, with 0 points assigned otherwise. In the end, the overall score was computed by summing the individual scores for each food component for each participant, yielding a score that ranged from 0 (low adherence) to 14 (high adherence) points.

The PDI is a measure of adherence to an overall plant-based diet, while hPDI and uPDI are measures of adherence to a healthy and unhealthy plant-based diets, respectively.[12] The scoring system is based on the classification of 18 food groups into three main categories: 7 healthy plant-based foods, 5 less healthy plant-based foods, and 6 animal-based foods. The healthy food groups consist of whole grains, fruits, vegetables, nuts, legumes, vegetable oils, and tea and coffee. Conversely, the less healthy food groups include fruit juices, refined grains, potatoes, sugar-sweetened beverages, and sweets and desserts. The animal food groups encompass animal fats, dairy products, eggs, fish and seafood, poultry and red meat, and miscellaneous animal-based foods. Each food group is ranked into quintiles and assigned positive or reverse scores. For positive scores, participants in the highest quintile of a food group receive a score of 5, and the score decreases by 1 for each subsequent quintile until participants below the lowest quintile receive a score of 1. The reverse scoring pattern is applied to the food groups with reverse scores. To create the PDI, positive scores are given to plant food groups, while animal food groups receive reverse scores. For the hPDI, positive scores are assigned to healthy plant food groups, while less healthy plant food groups and animal food groups receive reverse scores. Lastly, the uPDI assigns positive scores to less healthy plant food groups, while healthy plant food groups and animal food groups receive reverse scores. The individual scores for each of the 18 food groups are summed to obtain the indices, which range from 18 to 90 points. Higher scores on these indices indicate a lower intake of animal-based foods.

The Dietary Inflammatory Index (DII) is a comprehensive tool developed to assess the inflammatory potential of an individual's diet.[13] The construction of the DII involved an extensive literature review, where 1,943 articles were examined, focusing on 45 specific food parameters. The authors of the DII evaluated the impact of these dietary components on six key inflammatory biomarkers: IL-1β, IL-4, IL-6, IL-10, TNF-α, and CRP. Each food parameter was assigned a score based on its effect on the inflammatory biomarkers. A score of +1 indicated an increase in inflammation, -1 represented a decrease, and 0 indicated no effect. To establish a reference point, the DII authors compiled a "global standard mean" by averaging the intake levels of each DII component across 11 different datasets from various countries worldwide (USA, Australia, the Kingdom of Bahrain, Denmark, India, Japan, New Zealand, Taiwan, South Korea, Mexico, and the United Kingdom). The standard deviation was also calculated to provide context for the scores. The DII calculated for the present study consisted of 27 dietary components including anti- and pro-inflammatory nutrients and foods. The dietary components are categorized as anti-inflammatory: alcohol, monounsaturated fat, polyunsaturated fat, omega-3, omega-6, dietary fiber, beta carotene, folic acid, magnesium, thiamin, riboflavin, niacin, zinc, vitamins B6, A, C, D, E, green/black tea; and pro-inflammatory: total energy intake, carbohydrates, protein, total fat, saturated fat, cholesterol, vitamin B12 and iron and. A total of 18 components of the Shivappa et al. DII (i.e., turmeric, thyme/oregano, rosemary, eugenol, ginger, pepper, garlic, onion, caffeine, selenium, trans fat, isoflavones, flavan-3-ol, flavones, flavonols, flavonones, anthocyanidins, and saffron) were not available in our FFQ.[13] To calculate the DII score for each participant, their dietary intake of each DII component was compared to the global standard using Z-scores. The Z-score was obtained by subtracting the standard mean from the reported intake and dividing it by the standard deviation. These Z-scores were then transformed into centered percentile scores. To achieve a symmetrical distribution centered around 0, the percentile scores were doubled and '1' was subtracted. This transformation ensured that the values ranged from -1 (maximally anti-inflammatory) to +1 (maximally pro-inflammatory). The centered percentile values were subsequently multiplied by the overall pro- and anti-inflammatory effect score for each dietary component. The resulting overall DII score ranged from -4.37 to +4.56, with higher scores indicating a more pro-inflammatory dietary pattern, while lower DII scores represented a more anti-inflammatory diet.

**DNA methylation quantification**

Genomic DNA was extracted from buffy coat fractions of anti-coagulated blood samples using the Chemagic DNA buffy coat kit (PerkinElmer, Germany) with the Chemagic Magnetic Separation Module 1 and Chemagic Prime 8 Automated Workstation. Subsequently, the extracted DNA was bisulfite converted using the EZ-96DNA Methylation-LightningTMMagPrep from Zymo, following the manufacturer's instructions. DNA methylation levels were measured by Illumina iScan with Illumina's Human MethylationEPIC BeadChip, which measures approximately 850,000 CpG sites across the genome. Each probe's methylation level was determined as a beta value, representing the fractional level of DNA methylation at that specific probe. To ensure data quality, both sample-level and probe-level quality control procedures were performed using the 'minfi' package in R.[14] Samples were excluded from the analysis if they had sex mismatch (n= 9) or a missing rate exceeding 1% across all probes (n = 1). Similarly, probes with a missing rate higher than 1% (with a detection p-value greater than 0.01) across all participants were also excluded, following previously published recommendation guidelines for analyzing methylation data.[15]

**Gene expression profiling**

The gene expression profiling was performed on 750 ng of isolated total RNA. The RNA integrity and quantity were checked using TapeStation 4200 (Agilent) instrument and the NGS libraries for total RNA sequencing were generated through TruSeq stranded total RNA kit with Ribo-Zero Globin (Illumina). After checking the library size distribution (D1000 assay on TapeStation 4200, Agilent) and quantification (Qubit HS dsDNA assay, Invitrogen), the libraries were clustered at a final 250 pM concentration. The sequencing was performed in paired-end mode (2*50 cycles) on a NovaSeq6000 instrument (Illumina) using S2 v1 chemistry in XP mode, followed by demultiplexing using bcl2fastq2 v2.20. The reads sequence quality was assessed through FastQC v0.11.9 tool and the sequencing reads were aligned to the human reference genome GRCh38.p13 provided by Ensembl using STAR v2.7.1.[16] The count matrix was generated with STAR –quantMode GeneCounts using the human gene annotation version GRCh38.101.

We kept for subsequent analyses genes with an overall mean expression greater than 15 reads and expressed in at least 5% of the participants. Finally, the varianceStabilizingTransformation function from DESeq2 v1.30.1 R package was applied to normalize for library size and to log-transform the raw data.

**References**

1. Nöthlings, U., Hoffmann, K., Bergmann, M. M. & Boeing, H. Fitting Portion Sizes in a Self-Administered Food Frequency Questionnaire1,2. *The Journal of Nutrition* **137**, 2781–2786 (2007).
2. Richter, R., Tavares, J. F., Miloschewski, A., Breteler, M. M. B. & Mukherjee, S. ImputeBench: Benchmarking Single Imputation Methods. 2025.02.02.25321536 Preprint at https://doi.org/10.1101/2025.02.02.25321536 (2025).
3. BLS. https://www.blsdb.de/.
4. Slimani, N. *et al.* Structure of the standardized computerized 24-h diet recall interview used as reference method in the 22 centers participating in the EPIC project. *Computer Methods and Programs in Biomedicine* **58**, 251–266 (1999).
5. Trichopoulou, A., Costacou, T., Bamia, C. & Trichopoulos, D. Adherence to a Mediterranean diet and survival in a Greek population. *N Engl J Med* **348**, 2599–2608 (2003).
6. Fung, T. T. *et al.* Adherence to a DASH-style diet and risk of coronary heart disease and stroke in women. *Arch Intern Med* **168**, 713–720 (2008).
7. Morris, M. C. *et al.* MIND diet slows cognitive decline with aging. *Alzheimers Dement* **11**, 1015–1022 (2015).
8. Chiuve, S. E. *et al.* Alternative Dietary Indices Both Strongly Predict Risk of Chronic Disease123. *J Nutr* **142**, 1009–1018 (2012).
9. Galbete, C. *et al.* Nordic diet, Mediterranean diet, and the risk of chronic diseases: the EPIC-Potsdam study. *BMC Medicine* **16**, 99 (2018).
10. Willett, W. *et al.* Food in the Anthropocene: the EAT–Lancet Commission on healthy diets from sustainable food systems. *The Lancet* **393**, 447–492 (2019).
11. Knuppel, A., Papier, K., Key, T. J. & Travis, R. C. EAT-Lancet score and major health outcomes: the EPIC-Oxford study. *Lancet* **394**, 213–214 (2019).
12. Satija, A. *et al.* Healthful and unhealthful plant-based diets and the risk of coronary heart disease in US adults. *J Am Coll Cardiol* **70**, 411–422 (2017).
13. Shivappa, N., Steck, S. E., Hurley, T. G., Hussey, J. R. & Hébert, J. R. Designing and developing a literature-derived, population-based dietary inflammatory index. *Public Health Nutr* **17**, 1689–1696 (2014).
14. Fortin, J.-P., Triche, T. J., Jr & Hansen, K. D. Preprocessing, normalization and integration of the Illumina HumanMethylationEPIC array with minfi. *Bioinformatics* **33**, 558–560 (2017).

15. Wu, M. C. & Kuan, P.-F. A Guide to Illumina BeadChip Data Analysis. in *DNA Methylation Protocols* (ed. Tost, J.) 303–330 (Springer, New York, NY, 2018). doi:10.1007/978-1-4939-7481-8_16.
16. Dobin, A. *et al.* STAR: ultrafast universal RNA-seq aligner. *Bioinformatics* **29**, 15–21 (2013).
